# Evaluation of the accuracy, ease of use and limit of detection of novel, rapid, antigen-detecting point-of-care diagnostics for *SARS-CoV-2*

**DOI:** 10.1101/2020.10.01.20203836

**Authors:** L.J. Krüger, M. Gaeddert, L. Köppel, L. E. Brümmer, C. Gottschalk, I.B. Miranda, P. Schnitzler, H.G. Kräusslich, A.K. Lindner, O. Nikolai, F.P. Mockenhaupt, J. Seybold, V.M. Corman, C. Drosten, N.R. Pollock, A.I. Cubas-Atienzar, K. Kontogianni, A. Collins, A. H. Wright, B. Knorr, A. Welker, M. de Vos, J.A. Sacks, E.R. Adams, C.M. Denkinger, for the study team

## Abstract

**Background:** Reliable point-of-care (POC) diagnostics not requiring laboratory infrastructure could be a game changer in the COVID-19 pandemic, particularly in the Global South. We assessed performance, limit of detection and ease-of-use of three antigen-detecting, rapid POC tests (Ag-RDT) for *SARS-CoV-2*.

**Methods:** This prospective, multi-centre diagnostic accuracy study recruited participants suspected to have *SARS-CoV-2* in Germany and the UK. Paired nasopharyngeal swabs (NP) or NP and/or oropharyngeal swabs (OP) were collected from participants (one for clinical RT-PCR and one for Ag-RDT). Performance of each of three Ag-RDTs was compared to RT-PCR overall, and according to predefined subcategories e.g. cycle threshold (CT)-value, days from symptoms onset, etc. In addition, limited verification of the analytical limit-of-detection (LOD) was determined. To understand the usability a System Usability Scale (SUS) questionnaire and ease-of-use (EoU) assessment were performed.

**Results:** Between April 17^th^ and August 25^th^, 2020, 2417 participants were enrolled, with 70 (3.0%) testing positive by RT-PCR. The best-performing test (SD Biosensor, Inc. STANDARD Q) was 76×6% (95% Confidence Interval (CI) 62×8-86×4) sensitive and 99×3% (CI 98×6-99×6) specific. A sub-analysis showed all samples with RT-PCR CT-values <25 were detectable by STANDARD Q. The test was considered easy-to-use (SUS 86/100) and suitable for POC. Bioeasy and Coris showed specificity of 93×1% (CI 91×0%-94×8%) and 95×8% (CI 93×4%-97×4%), respectively, not meeting the predefined target of ≥98%.

**Conclusion:** There is large variability in performance of Ag-RDT with SD Biosensor showing promise. Given the usability at POC, this test is likely to have impact despite imperfect sensitivity; however further research and modelling are needed.

## Introduction

As of 23^rd^ of September 2020, *SARS-CoV-2* has affected the entire globe with over 30 million confirmed infections worldwide, leading to almost one million fatalities. ^1^ Already early in the pandemic, the World Health Organization (WHO) highlighted the critical importance of testing for epidemic control. ^2^ Reverse transcription polymerase chain reaction (RT-PCR) is the current gold standard for the detection of *SARS-CoV-2*. ^3^ Specialized laboratories for RT-PCR testing are available in high-and low-resource settings. ^4^ However, global shortages of test reagents and limited infrastructure and human resource capacity, have resulted in unavailability of testing or long turnaround times, especially in the Global South. To combat transmission, fast, easy-to-use, accurate, and point-of-care (POC) diagnostic tools are needed on a large scale.

Detection of pathogen-derived antigens on lateral flow assays is a proven concept with excellent performance, e.g. for malaria in blood. ^5^ This method enables rapid, instrument-free, and low-cost POC testing. In respiratory samples, however (e.g. for diagnosis of influenza), performance has been variable. ^6^

Since the onset of the pandemic, numerous companies have been working on bringing a rapid *SARS-CoV-2* antigen-detection test (Ag-RDT) to the market. ^7^ Interest and funding support are substantial ^8^, and the USA, India and Japan, among other countries have approved their use. ^9-11^ WHO has recently released interim guidance that the use of Ag-RDTs that meet at least 80% sensitivity and 97% specificity may be considered to diagnose active *SARS-CoV-2* infection “where nucleic acid amplification technology (NAAT) is unavailable or where prolonged turnaround times preclude clinical testing.”. ^12,13^ Few peer-reviewed studies evaluating these novel assays have been published with only one small study, to our knowledge, performed independently of the manufacturer. ^14,15^

Here we present a manufacturer-independent validation of three novel, antigen-detecting, rapid POC diagnostics for *SARS-CoV-2*. Specifically, we performed a verification of analytical sensitivity, a large multi-centre clinical accuracy study, and an ease-of-use (EoU)assessment. The work was conducted in partnership with the Foundation of Innovative New Diagnostics (FIND) and has informed a WHO recommendation. ^16^ FIND is the WHO collaborating centre for Coronavirus disease 2019 (COVID-19) diagnostics.

## Methods

### Ethic statement

The study protocol was approved by the ethical review committee at the Heidelberg University Hospital for the study sites in Heidelberg and Berlin (Registration number S-180/2020), and by the NHS review board, IRAS number 282147, for the study in Liverpool, UK. Each participant provided written informed consent.

### Tests evaluated

Three novel rapid antigen-based diagnostics were assessed

(a) Bioeasy 2019-nCoV Ag Fluorescence Rapid Test Kit (Time-Resolved Fluorescence). ^17^ (Shenzhen Bioeasy Biotechnology Co. Ltd., Guangdong Province, China; henceforth called Bioeasy), (b) COVID-19 Ag Respi-Strip ^18^ (Coris Bioconcept, Gembloux, Belgium; henceforth called Coris) and (c) STANDARD Q COVID-19 Ag Test ^19^ (SD Biosensor, Inc. Gyeonggi-do, Korea; henceforth called SD Biosensor).

All assays utilize the lateral flow assay principle for the detection of viral antigens. Both Coris and SD Biosensor use colloidal gold conjugated to antibodies to enable a colour change and read-out with the naked eye, while Bioeasy uses fluorescent readout and thus requires the use of a proprietary reader device. Coris is designed as a dipstick, whereas Bioeasy and SD Biosensor use a cassette-based assay format. For Coris and SD Biosensor the manufacturers’ instructions for use (IFU) were followed; except for Bioeasy, the IFU was followed except the developer requested for pipettes to be used to transfer adequate quantities of liquid (in the IFU no pipette is needed and a nozzle is provided). ^17^ Bioeasy and SD Biosensor include swabs within their test kits for sample collection, while for Coris, a commercially available swab was used: the eSwab system from Copan Diagnostics (Murrieta, CA, USA), which contains 1ml of Amies solution and was approved for use by Coris. A prespecified quantity of proprietary buffer solution was applied to the lateral flow cassette for Bioeasy and SD Biosensor and into the test tube with the dipstick for Coris as per the respective IFU. The readout was done within the recommended time for each Ag-RDT (10 minutes for Bioeasy, 15 minutes for Coris and 15 to 30 minutes for SD Biosensor).

### Analytical study

A limited verification of the analytical sensitivity (lower limit of detection) was performed using standardized, quantified viral cultures.

The *SARS-CoV-2* isolate REMRQ0001/Human/2020/Liverpool was propagated in Vero E6 cells (C1008; African green monkey kidney cells). Cells were maintained in Dulbecco’s minimal essential medium (DMEM) containing 10% foetal bovine serum (FBS) and 0×05 mg/mL gentamycin at 37°C with 5% CO2 and FBS concentration was reduced to 4% for viral propagation.

Ten-fold serial dilutions of virus stock starting from 1×0 × 10^6^ Plaque forming units (PFU)/ml were made using culture media as a diluent (DMEM + 2% FBS % + 0.05 mg/ml gentamycin). Two-fold dilutions were made between 1×0 × 10^4^ to 1×0 × 10^2^. Lateral flow tests were performed in triplicate. For SD Biosensor and Bioeasy, the swab included in each individual kit was immersed into the virus dilutions and spiked (soaked). Subsequently, the test was performed using the spiked swab following the manufacturer’s instructions. For Coris, the eSwab from Copan was spiked with serial dilutions of virus and then added to the 1 ml of Amies solution contained in the swab tube. 100μl of each of the sample dilutions were added to the test tube with the dipstick and was read-out as per manufacturer’s instructions.

Coris and SD Biosensor tests were read by two operators, each blinded to the result of the other. A third operator read any discrepant tests and final result was agreed (2/3). The Bioeasy result is a digital readout, and the first operator recorded the result, which was checked by the second operator, and printed out directly from the machine.

### Clinical diagnostic accuracy study

#### Study design and participants

The primary objective of the clinical multicentre diagnostic accuracy study was to estimate the sensitivity and specificity of a single antigen test compared to a single PCR test as the reference method.

Participant recruitment and sample collection was conducted at three sites: (1) A drive-in testing station in Heidelberg, Germany; (2) a clinical ambulatory testing facility in Berlin, Germany and (3) secondary care facilities at Liverpool University Hospital Foundation Trust, Royal and Aintree (LUHFT), Liverpool, United Kingdom. Participants screened for the study were adults (age ≥18 years) identified as at risk for *SARS CoV-2* infection according to the local department of public health based on exposure to a *SARS CoV-2* positive person, suggestive symptoms, or travel to a high risk area. Participants were excluded if they had previously been diagnosed with *SARS CoV-2* or if their command of either English or German was insufficient to give informed consent for participation.

### Study procedures

#### Antigen-detecting testing

After obtaining written consent, participants underwent a first swab for testing with RT-PCR, which was taken either as (i) a nasopharyngeal (NP) swab or (ii) oropharyngeal (OP) swab (if clinically contra-indicated to do a NP swab (e.g. because of risk of bleeding)) in Heidelberg or (iii) combined as NP/OP swab in Berlin and Liverpool as per institutional recommendations (with OP done first, followed by NP with the same swab). Subsequently, participants underwent collection of the same type of swab again for Ag-RDT.

Ag-RDTs were performed at the sample collection site in Heidelberg and Berlin. In Liverpool samples were transported on ice to a category 3 facility and then processed. For on-site testing, separate areas were maintained for clean material (without sample) and ‘unclean’ areas of benches (material with sample) to avoid cross-contamination. Disinfection was performed and gloves exchanged after each sample. Negative control testing with buffer without sample was performed regularly to capture possible contamination. For the Ag-RDTs with visual read-out (Coris and SD Biosensor), the results were interpreted by two operators, each blinded to the result of the other. If a discrepant result was obtained, both operators re-read the result and agreed on a final result. Invalid results were repeated once using the remaining buffer according to the respective IFUs.

#### RT-PCR testing

The swab (eSwab, Copan) for RT-PCR testing was sent to the referral laboratory in the provided Amies solution. The RT-PCR assays were performed according to routine procedures at the referral laboratory. In Heidelberg, the *SARS CoV-2* assay from TibMolbiol (Berlin, Germany), the Allplex *SARS-CoV-2* Assay from Seegene (Seoul, South Korea) or the Abbott (Illinois, US) RealTime 2019-nCoV assay was performed. In Berlin the Roche Cobas *SARS CoV-2* assay (Pleasanton, CA United States) on the Cobas^®^ 6800 or 8800 system or the *SARS CoV-2* assay from TibMolbiol (Berlin, Germany) was performed. In Liverpool, the Genesig^®^ Real-Time Coronavirus COVID-19 PCR assay (Genesig, UK) was performed. Assays were calibrated with the E-gene assay as described before. ^3,19^ Only samples that showed a signal above the threshold in the relevant RT-PCR target regions for each assay were considered to be positive. Staff performing the Ag-RDTs were blinded to results of RT-PCR tests and vice versa.

After completion of testing, the participants in Heidelberg were contacted for an interview focusing on symptoms and co-morbidities via telephone or email according to the indicated preference on site. In Berlin and Liverpool participants were interviewed directly on-site after obtaining the consent (questionnaire available in the supplement material Section (B)).

All data were captured in a dedicated database utilizing Research Electronic Data Capture (REDCap^™^, www.prorect-redcap.org) hosted at the Heidelberg University Hospital or Liverpool School of Tropical Medicine (LSTM). REDCap^™^ is a secure, web-based application, which provides audit trails for tracking data manipulation and export procedures. The protocol is provided in the supplement material Section (C).

### System Usability Scale and Ease of Use Assessment

To understand the usability of the three Ag-RDTs, a standardized System Usability Scale (SUS) ^20^ questionnaire and a ease-of-use assessment (EoU), specifically developed for the study, were performed. The questionnaire can be found in the supplement material Section (D). The laboratory personnel from each study site (at least 3 operators per test) were invited to complete the surveys separately for each assay. The complete EoU survey can be found in the supplement material Section (D). For the interpretation of the SUS, it can be said that a SUS score above 68 would be considered above average and anything below the score of 68 is below average. ^20^

### Statistical Analysis

In the analytical assessment, the limit of detection (LOD) was determined as the lowest concentration for which all three replicates scored positive.

In the clinical accuracy study, the sensitivity, specificity, positive predictive value (PPV), and negative predictive value (NPV) of the Ag-RDTs with 95% confidence intervals (CIs) were estimated by comparing the Ag-RDT result to the gold standard, RT-PCR, on the same day from the same participant (statistical analysis plan available upon request). A sub-analysis was performed by CT-value. ^21-23^ Furthermore, a conversion of CT-values for RT-PCR tests into viral-load was performed by calibrating the test systems using quantified specific in vitro-transcribed RNA. ^24^

A sample size of 1000 participants per Ag-RDT under evaluation was targeted but given the variable prevalence throughout the pandemic this target was left flexible. Interim analyses were performed at predefined sample sizes (25%, 50%) and an evaluation of an Ag-RDT was stopped if the predefined performance criterion for specificity (≥98%, considering the upper bound of the confidence interval) was not met. ^25^ Invalid Ag-RDT results were reported separately, whereas participants with invalid RT-PCR results were excluded from the analysis. Inter-operator variability was assessed for tests read with the naked eye. The analysis was conducted with the statistical software R (R Foundation for Statistical Computing, Vienna, Austria).

For the usability assessment, the SUS score was calculated taking the mean of all answers given for each test. For the EoU assessment, verbal answers were transferred into numerical values. Taking the mean across values, all answers given for each test were summarized into a heat map. Both SUS and Ease of Use assessment were analysed with Microsoft Excel.

### Role of the funding source

The study was supported by FIND, Heidelberg University Hospital and Charité – University Hospital internal funds. Pfizer funded the clinical team in Liverpool, UK. FIND provided input on the study design, and data analysis in collaboration with the rest of the study team. The other external funders of the study had no role in study design, data collection, or data analysis. The corresponding author had full access to all the data in the study and had final responsibility for the decision to submit for publication.

## Results

### Analytical sensitivity and exclusivity

SD Biosensor was positive in all replicates at 5×0 × 10^3^ PFU/ml, and only one out of three replicates at 1×0 × 10^3^ PFU/ml, which is approximately ten-fold higher than the supplier-reported LOD of 3×06 × 10^2.2^ TCID50/ml as per ATCC.org. ^19^ The Bioeasy assay was positive on all three replicates at 5×0 × 10^3^ PFU/ml (approximately 7×14 × 10^3^ TCID50/ml) and only one out of three replicates at 2×5 × 10^3^ PFU/ml (approximately 3×57 × 10^3^ TCID50/ml). Coris was positive on all three replicates at 1×0 × 10^4^ PFU/ml (approximately 1×43 × 10^4^ TCID50/ml), which is two-fold higher than the supplier-reported LOD of 7×14 × 10^3^ TCID50/ml. ^26^ Coris was positive on only one of three replicates at 5×0 × 10^3^ PFU/ml (approximately 7×14 × 10^3^ TCID50/ml).

### Clinical diagnostic accuracy study

Between April 17^th^ and August 25^th^ 2020, 3160 participants with presumed *SARS-CoV-2* infections were invited to participate in the study. In total, 2417 met inclusion criteria, provided consent and were included in the analysis (Figure 1). Of those, 1263 participants were tested using SD Biosensor from July 20^th^ to July 31^st^ 2020 in Heidelberg (N=334), from June 3^rd^ to July 31^st^ 2020 (N=910) in Berlin, and May 20^th^ to June 12^th^ 2020 (N=19) in Liverpool. Separately, 729 participants were tested using Bioeasy, 506 of whom were enrolled in Heidelberg between April 17^th^ and May 3^rd^ 2020, and 223 in Berlin between May 14^th^ and June 3^rd^ 2020. 425 enrolled participants were tested using Coris: 286 in Heidelberg from 11^th^ to 25^th^ May 2020; 105 in Berlin from 19^th^ to 25^th^ August 2020; and 34 in Liverpool from May 12^th^ to June 12^th^ 2020. The variance in sample size is explained by the fact that both the Coris and Bioeasy evaluations were aborted after interim analysis indicated that specificity did not meet predefined performance criteria, and after a work-up for reasons for the false-positive results was completed.

**Figure 1.**
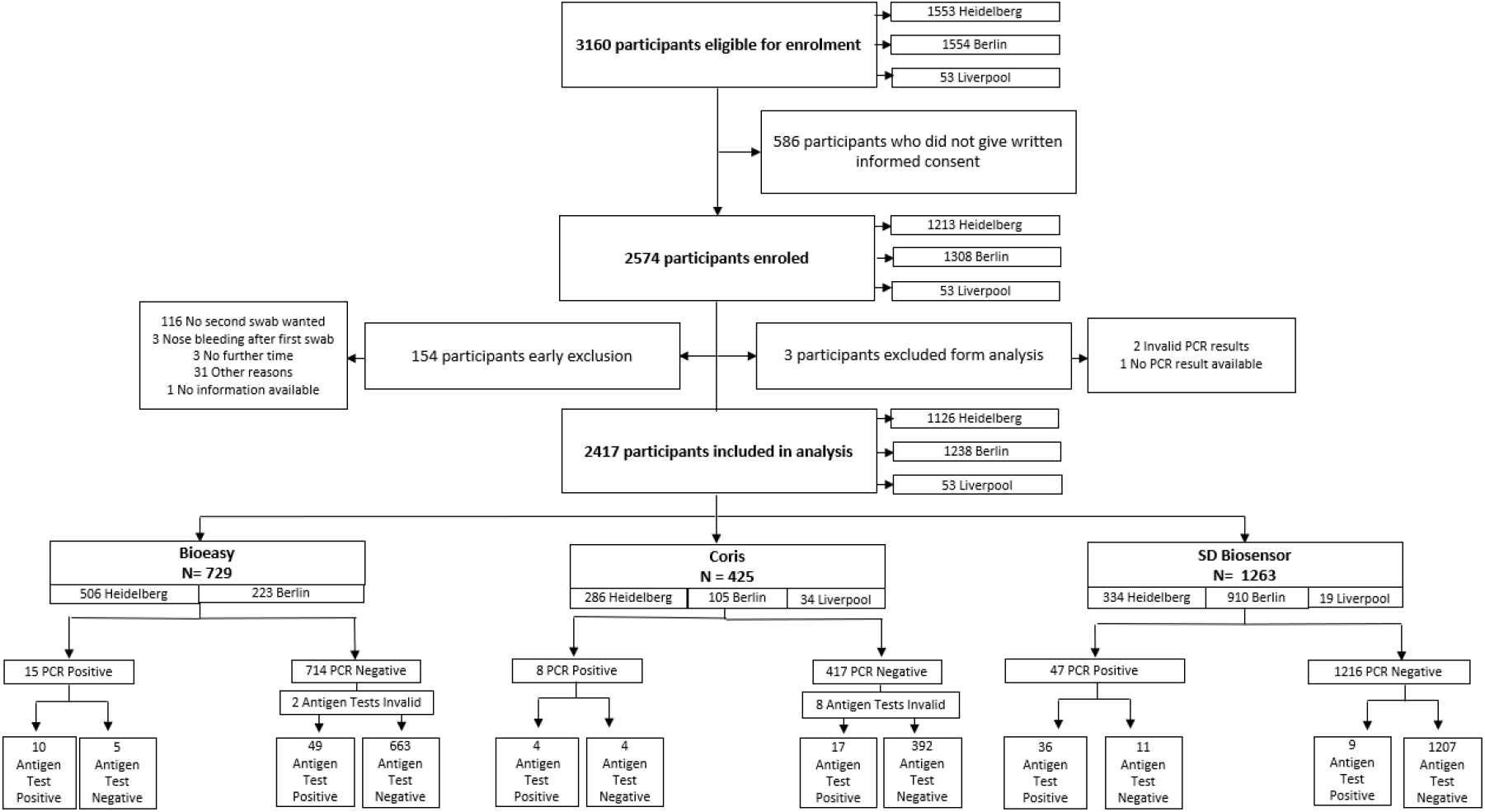
Study Flow.

We present clinical and demographic characteristics overall and by Ag-RDT in Table 1 (Supplement Tables 2, 3 and 4 show study characteristics by study site). The average age of participants presenting for testing was 40×4 years (Standard Deviation [SD] 14×3) with 52×8% being female and 35×9% having comorbidities. On the day of testing, 80×7% of participants had one or more symptoms consistent with COVID-19 (questionnaire in the supplement material Section (B)), with most (1868 (79×3%)) reporting being only mildly ill and only 6 (0×3%) stating that they are completely bedridden due to illness (a more detailed list of symptoms and comorbidities is provided in the supplement Table 5). Average duration of symptoms at the time of swab collection/testing was 5×0 days (SD 9×6 days). Of the 2407 analysed participants, 70 (2×9%) tested positive by RT-PCR, of which 30 (42×9%) had CT values of <25. A description of symptoms for all participants tested positive by RT-PCR is provided in supplement Table 5.

**Table 1.**
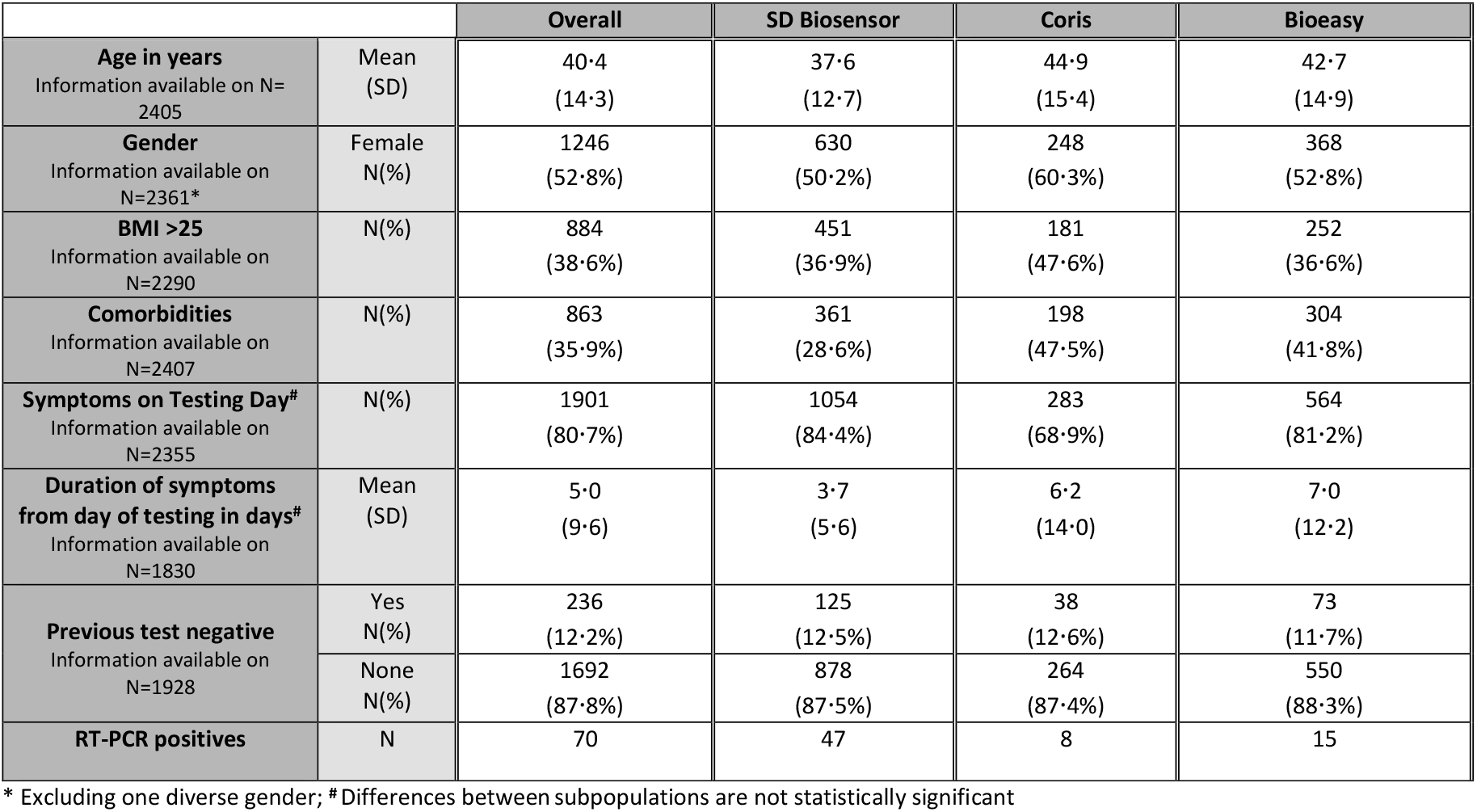
Study population characteristics.

|  |  | Overall | SD Biosensor | Coris | Bioeasy |
| --- | --- | --- | --- | --- | --- |
| **Age in years**<br>Information available on N=2405 | Mean (SD) | 40·4<br>(14·3) | 37·6<br>(12·7) | 44·9<br>(15·4) | 42·7<br>(14·9) |
| **Gender**<br>Information available on N=2361* | Female<br>N(%) | 1246<br>(52·8%) | 630<br>(50·2%) | 248<br>(60·3%) | 368<br>(52·8%) |
| **BMI >25**<br>Information available on N=2290 | N(%) | 884<br>(38·6%) | 451<br>(36·9%) | 181<br>(47·6%) | 252<br>(36·6%) |
| **Comorbidities**<br>Information available on N=2407 | N(%) | 863<br>(35·9%) | 361<br>(28·6%) | 198<br>(47·5%) | 304<br>(41·8%) |
| **Symptoms on Testing Day<sup>#</sup>**<br>Information available on N=2355 | N(%) | 1901<br>(80·7%) | 1054<br>(84·4%) | 283<br>(68·9%) | 564<br>(81·2%) |
| **Duration of symptoms from day of testing in days<sup>#</sup>**<br>Information available on N=1830 | Mean (SD) | 5·0<br>(9·6) | 3·7<br>(5·6) | 6·2<br>(14·0) | 7·0<br>(12·2) |
| **Previous test negative**<br>Information available on N=1928 | Yes<br>N(%) | 236<br>(12·2%) | 125<br>(12·5%) | 38<br>(12·6%) | 73<br>(11·7%) |
|  | None<br>N(%) | 1692<br>(87·8%) | 878<br>(87·5%) | 264<br>(87·4%) | 550<br>(88·3%) |
| **RT-PCR positives** | N | 70 | 47 | 8 | 15 |

\* Excluding one diverse gender; # Differences between subpopulations are not statistically significant

The best performing test among the three was SD Biosensor with a sensitivity of 76×6% (36 out of 47 positives detected; CI 62×8-86×4) and specificity of 99×3% (CI 98×6-99×6) (Table 2). The sensitivity was 100% (CI 82×4-100) for the 18 samples with CT-values <25, while for those with CT ≥25, only 18 out of 29 scored positive (62×1%; CI 44×0-77×3) (Table 3). Corresponding viral load to CT values of RT-PCR positive results are provided in the supplement Table 6. A sub-analysis of sensitivity of SD Biosensor by sampling strategy suggested that a NP swab alone might be less effective than a combined NP/OP swab, however confidence intervals were overlapping (Table 4). No difference was seen by duration of symptoms (one vs two weeks). A trend was observed for increased sensitivity by disease severity, although again confidence intervals were overlapping (58×8% [36×0-78×4], 85×7% [65×4-95×0], 100% [51×0-100] for normal activity, mild and severe illness, respectively). No invalid results occurred. Two test results were excluded due to handling issues (read-out after time limit). Inter-operator reliability was near perfect with a kappa of 0×99. Overall, the usability was considered very favourable, with a SUS score of 86/100, and the only challenges flagged by the EoU were occasional difficulties when interpreting the test result (i.e. intensity of band; Figure 2).

**Table 2.**
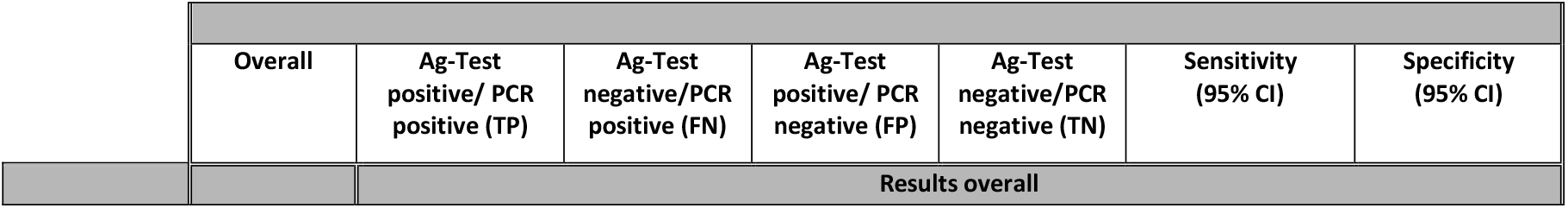
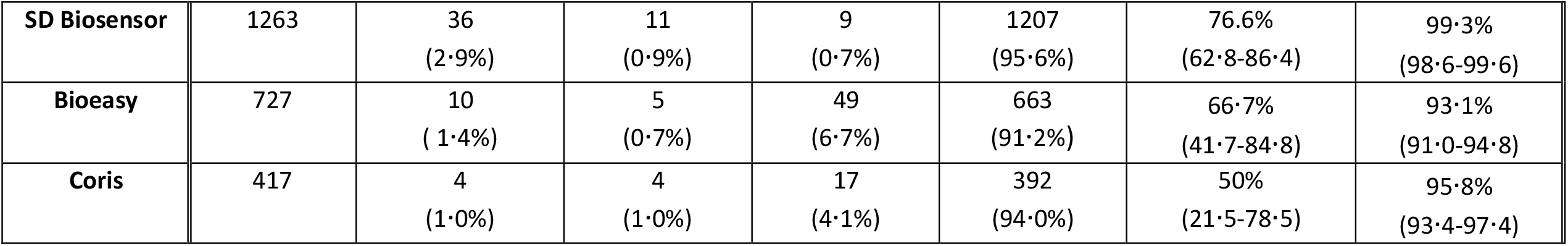
Antigen test performance overall.

**Table 3.** Subanalysis for antigen test performance in different CT-value categories for only RT-PCR positive results.

|  | Overall | Ag-Test positive | Ag-Test negative | Sensitivity<br>(95% CI) |
| --- | --- | --- | --- | --- |
| **CT value PCR <25** |  |  |  |  |
| **SD Biosensor** | 18<br>(60%) | 18<br>(100%) | 0<br>(0%) | 100%<br>(82·4-100) |
| **Bioeasy** | 9<br>(30%) | 8<br>(88·9%) | 1<br>(11·1%) | 88·9%<br>(56·5-99·4) |
| **Coris** | 3<br>(10%) | 2<br>(66·7%) | 1<br>(33·3%) | 66·7%<br>(20·8-98·3) |
| **CT value PCR ≥25** |  |  |  |  |
| **SD Biosensor** | 29<br>(69%) | 18<br>(62·1%) | 11<br>(37·9%) | 62·1%<br>(44·0-77·3) |
| **Bioeasy** | 8<br>(19%) | 2<br>(25%) | 6<br>(75%) | 33·33%<br>(9·7-70·0) |
| **Coris** | 5<br>(12%) | 2<br>(40%) | 3<br>(60%) | 40%<br>(11·8-76·9) |

**Table 4.** Subgroup Analysis for SD Biosensor, Inc. STANDARD Q.

|  | Overall | Ag-Test<br>positive/ PCR<br>positive | Ag-Test<br>negative/PCR<br>positive | Ag-Test<br>positive/ PCR<br>negative | Ag-Test<br>negative/PCR<br>negative | Sensitivity<br>(%, 95% CI) | Specificity<br>(%, 95% CI) |
| --- | --- | --- | --- | --- | --- | --- | --- |
| **Sampling strategy** |  |  |  |  |  |  |  |
| **NP swab**<br>Heidelberg | 299<br>(23·7%) | 4 | 3 | 6 | 286 | 57·1%<br>(25·0-84·1) | 97·9%<br>(95·6-99·1) |
| **OP swab**<br>Heidelberg | 35<br>(2·8%) | 0 | 0 | 0 | 35 | NA (no positive<br>PCR results) | 100%<br>(90·1-100) |
| **NP/OP swab\***<br>Berlin | 907<br>(71·8%) | 31 | 8 | 3 | 865 | 79·5%<br>(64·5-89·2) | 99·7%<br>(99·0-99·9) |
| **NP/OP swab**<br>Liverpool | 19<br>(1·5%) | 0 | 0 | 0 | 19 | NA (no positive<br>PCR results) | 100%<br>(83·2-100) |
| **Symptom duration – Information available for N=1018<sup>+</sup>** |  |  |  |  |  |  |  |
| **0-7 days**<br>Overall | 949 (93·2%) | 28 | 7 | 7 | 907 | 80·0%<br>(64·1-90·0) | 99·2%<br>(98·4-99·6) |
| **8-14 days**<br>Overall | 58 (5·7%) | 4 | 0 | 0 | 54 | 100 %<br>(51·0-100) | 100%<br>(93·0-100) |
| **> 14 days**<br>Overall | 11 (1·1%) | 0 | 0 | 0 | 11 | NA (no positive<br>PCR results) | 100%<br>(74·1-100) |
| **Disease severity in 3. categories – Information available for N=1193** |  |  |  |  |  |  |  |
| **Normal activity possible**<br>Overall | 495 (41·5%) | 10 | 7 | 4 | 474 | 58·8%<br>(36·0-78·4) | 99·2%<br>(97·9-99·7) |
| **Light restrictions**<br>and<br>**Able to walk**<br>Overall | 673 (56·4%) | 18 | 3 | 5 | 647 | 85·7%<br>(65·4-95·0) | 99·2%<br>(98·2-99·7) |
| **Limited self-sufficiency**<br>and<br>**Completely need of care**<br>Overall | 25 (2·1%) | 4 | 0 | 0 | 21 | 100%<br>(51·0-100) | 100%<br>(84·5-100) |

\* 3 participants in Berlin did not receive the combined swab; <sup>+</sup> No information from N=8 PCR positives out of N=47

**Figure 2.**
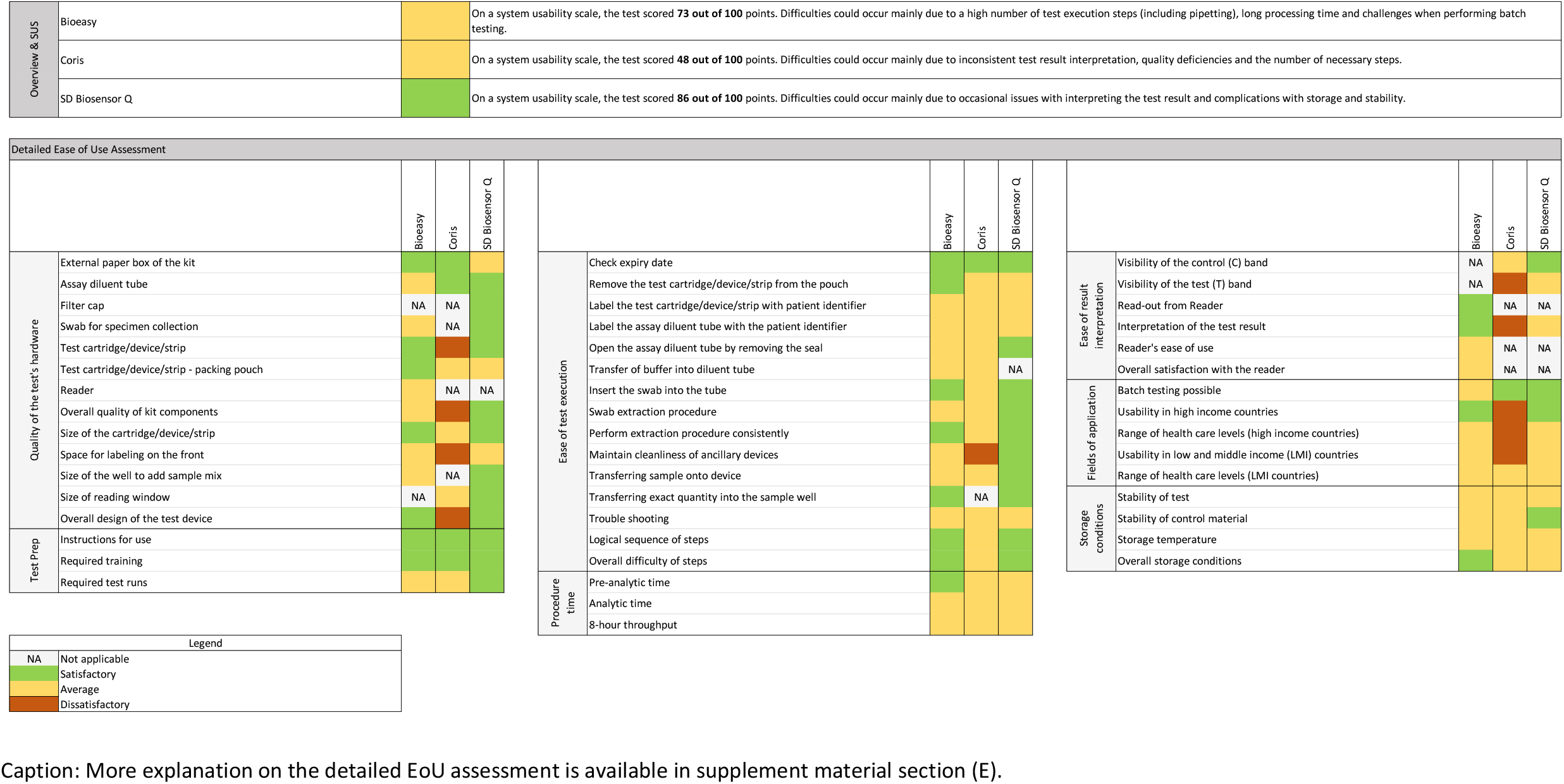
System Usability Score and Ease of Use assessment results. Caption: More explanation on the detailed EoU assessment is available in supplement material section (E).

Bioeasy had a sensitivity of 66×7% (10 out of 15 detected; CI 41×7-84×9) and specificity of 93×1% (49 false-positives; CI 91×0-94×8) (Table 2.). The sensitivity was 88×9% (8/9 detected; CI 56×5-99×4) for samples with CT-values <25, while for those with CT ≥25 only 2 out of 6 tested positive (33×3%; CI 9×7-70×0). Further sub-analyses were not performed for sensitivity because of the limited number of positive samples. Two invalid results occurred in 729 tests (0×3%). Overall, the usability was considered favourably with a SUS score of 73/100. The EoU identified a high number of test execution steps (including precision pipetting) which resulted in challenges when performing multiple tests at the same time possibly hindering the test’s wide-spread use (Figure 2).

Coris showed a sensitivity of 50% (4 out of 8; CI 21×5-78×5) and specificity of 95×8% (CI 93×4-97×4). Two out of the three samples with CT <25 were detected, while for the five specimens with CT ≥25, sensitivity was 40%. Further sub-analyses were not performed for sensitivity because of the limited number of positive samples. The low specificity determined the stop of the study as the upper margin of 95% CI was below 98%. Inter-operator reliability was very good with a kappa of 0×95. Invalid tests occurred on 8 occasions (1×9%) either due to the control line not being visible or due to smearing of the readout window. Four results were excluded due to handling issues (read-out after time limit). Overall, the usability was considered unfavourable with a SUS score of 48/100 and the EoU pointed towards challenges due to inconsistent test result interpretation (often only very faint lines visible) and deficiencies in both the test kit quality and design (Figure 2).

An investigation of the false-positives was performed for all assays by (1) testing negative buffer (without swab) interspersed with patient samples; (2) repeating RT-PCR from Ag-RDT buffer/swab sample for a subset of false-positive Ag-RDT samples (where feasible from available quantity of buffer); (3) having the sampling performed by the same person for Ag-RDT and PCR test (to limit variability in sampling); and (4) contacting participants again and retesting with Ag-RDT and RT-PCR if they had persistent symptoms or COVID-19 cases in their surroundings. None of this work-up yielded any evidence of cross-contamination, limited RT-PCR sensitivity or sampling error, which makes cross-reactivity most likely as the reason for the false-positives (target of cross-reactivity unknown).

## Discussion

Results of our multi-centre diagnostic accuracy study and analytical assessment show that the SD Biosensor, Inc. STANDARD Q Ag-RDT COVID-19 Ag test can reach excellent sensitivity in high-viral load infections, and good performance in the cohort overall 76×6% (CI 62×8-86×4) with near-perfect specificity (99×3%; CI 98×6-99×6). These results hold promise for the performance of antigen-based diagnosis and the control of the pandemic, particularly in low-resource settings.

*SARS CoV-2* is a virus with a low dispersion factor, which indicates that a small number of infected individuals are responsible for a large amount of the transmission - the so-called ‘super spreaders’. ^27^ A super-spreader is likely to both have a high viral load, and the occasion to transmit to many people due to aggregation of people in confined spaces. ^28^ With a sensitivity of 100% for the SD Biosensor in infected persons with a high viral load, this Ag-RDT is likely to identify highly contagious individuals, which would more substantially decrease transmission than what would be expected from its imperfect overall sensitivity alone. Furthermore, the rapid turn-around time is likely to result in more rapid isolation of cases and effective contact tracing. The ease-of-use and POC-suitability can enable more frequent testing as compared to RT-PCR and thus, despite its limited sensitivity, the test is expected to have impact on pandemic control based on a recent modelling study. ^29^

Considering the test in low prevalence settings (prevalence 1%), a negative test would have a negative predictive value (NPV) of 99×8%. And even at a prevalence of 10%, 97×4% of negatively tested persons would not be infected. Such an NPV would be considered acceptable outside of high-risk transmission settings (e.g. in a hospital). The positive predictive value (PPV) would be 53% at 1% prevalence, ideally requiring RT-PCR confirmation. At 10% prevalence, the PPV increases to 92%. This suggests that several use-cases could be considered with substantial public health impact, but further implementation research is needed to better detail how the tests can be used to the highest benefit. Furthermore, stability only up to 30° Celsius and required storage conditions are a limitation for the use in the Global South, and should be considered carefully in such settings.

Our study also shows wide variability in performance between Ag-RDTs with respect to performance and ease-of-use, indicating the importance of manufacturer-independent validations. While our findings largely corroborate the performance reported in the IFU of SD Biosensor, for Coris and Bioeasy, they differed substantially in respect to specificity. For SD Biosensor, the lower overall performance compared to the results in the IFU could relate to the fact that for some patients in our study OP swabs were used. While this is not recommended by the manufacturer, the use of OP swabs when NP is contraindicated, reflects a more clinically realistic scenario. NP/OP swabs were used based on institutional recommendations in Berlin and Liverpool. While more proteases are expected in the saliva that can degrade antigen, there should not be an impact on the sensitivity of Ag-RDTs in our study, as swabs were processed immediately. Another aspect that could explain a lower senstivtiy is the population tested in the study. The population tested was likely representative of the spectrum of COVID-19, with less ill patients likely to have lower viral load at diagnosis and more likely Ag-RDT negative. A higher clinical sensitivity would be expected when only hospitalized patients were tested.^30^ For Bioeasy, our point estimate of specificity falls within the confidence interval of specificity of the only other independently published study, which had few RT-PCR negative cases. ^14^

Overall, our study had several strengths. The study confirmed the accuracy of the tests by two measures: analytical and clinical accuracy. The population enrolled for testing was representative of the pandemic observed in Europe with a broad spectrum of age group (initially older age group and with ongoing duration of the pandemic an increasingly younger age group presenting for testing) and a spread of clinical presentations (from asymptomatic with high-risk contacts to severely ill). We enrolled in two countries and although enrolment numbers in Liverpool were low, the results confirmed findings in Germany. Due to the wide-spread testing available particularly in Germany, the population tested is expected to be a representative spectrum of disease. The tests were performed at POC at two sites (Berlin and Heidelberg) thus mimicking the real-world challenges of POC testing. The comprehensive ease-of-use assessment with a standardized SUS-tool and a questionnaire, developed specifically for the study, captured the differences between the tests and highlighted important points for operationalization of the tests.

However, the study also has several limitations. First, the prevalence of positive cases decreased substantially over the course of the study from 10% among tested individuals in early April as documented by the local health department in Heidelberg, Germany, to 3% over the course of the study. In addition, our study assessed a preselected population based on the regulations and criteria from the local health department. Due to the nature of the pandemic, the selection criteria and patient population changed throughout the study as visible from the percentage of patients with symptoms and co-morbidities evaluated with the three Ag-RDTs, although differences between the study populations were not significant. In addition, our study did use OP swabs for a subset of patients as discussed above. Furthermore, the sub-analysis by CT-values has its limitations, as RT-PCR methods varied across sites with different genome targets, PCR instruments and reagents. This might cause misleading comparisons of CT-values between sites. ^23^ Therefore, we only performed an analysis of dichotomized CT-values and considered <25 as high viral load and ≥25 as low viral load. In addition, we translated CT-values into viral load, and determined the variability of the applied *SARS-CoV-2* assays in testing of standardized dilutions of virus to be limited, as shown before. ^21,22^

### Conclusion

Our results suggest that, though there is substantial variability between tests, at least one antigen-detection POC test meets WHO targets and is now recommended by the WHO. ^16^ Given the fast turn-around-time and ease-of-use at POC, as well as the high sensitivity in patients with significant viral load, the test is likely to have impact despite imperfect overall sensitivity; however, further implementation research, economic evaluation and modelling are needed. With the agreement of Roche and SD Biosensor, manufacturing scale-up and supply-chain solutions should guarantee availability of the test. ^31^

## Data Availability

The data will be available upon request.

## Notes

### Competing Interest Statement

The authors have declared no competing interest.

### Clinical Trial

German Clinical Trial Registry DRKS00021220

### Clinical Protocols

https://apps.who.int/trialsearch/Trial2.aspx?TrialID=DRKS00021220

### Funding Statement

The study was supported by FIND, Heidelberg University Hospital and Charite University Hospital internal funds. Pfizer funded the clinical team in Liverpool, UK. The external funders of the study had no role in study design, data collection, or data analysis. The corresponding author had full access to all the data in the study and had final responsibility for the decision to submit for publication.

### Author Declarations

The study protocol was approved by the ethical review committee at the University Hospital Heidelberg for the study sites in Heidelberg and Berlin (Registration number S-180/2020), and by the NHS review board, IRAS number 282147, for the study in Liverpool, UK. Each participant provided written informed consent.

