## Supplement for "Evaluation of the accuracy, ease of use and limit of detection of novel, rapid, antigen-detecting point-of-care diagnostics for *SARS-CoV-2*": Supplementary material_0.11_30.09.20.pdf

#### (A) Table 1: Study Team

| | |
|---|---|
| Department of Public Health Rhein Neckar Region, Heidelberg, Germany | Dr. K. Assaad, |
| | Dr. A. Fuhs |
| | C.Harter |
| | C,. Schulze |
| | G. Schmitt |
| Division of Clinical Tropical Medicine, Heidelberg University Hospital, Germany | Anja Klemmer |
| | Paula Gieser |
| | Maximilian Schirmer |
| | Sven Blobner |
| | Matthias Meinlschmidt |
| | Jana Schmehl |
| | Marta Machado |
| | Ann-Kathrin Huber |
| | Nick van der Hoeven |
| | Tim Starck |
| | Valerie Dürr |
| | Martina Fink |
| | Felicia Margono |
| | Kathinka Buchholz |
| | Jan Klaas Janßen |
| | Sofie Knoll |
| | Alexander Penning |
| | Salome Steinke |
| Institute of Tropical Medicine and International Health, Charité – Universitätsmedizin Berlin, Berlin, Germany | Mandy Kollatzsch |
| | Mia Wintel |
| | Franka Kausch |
| | Franziska Hommes |
| | Alisa Bölke |
| | Julian Bernhard |
| | Claudia Hülso |
| | Elisabeth Linzbach |
| Medical Directorate, Charité – Universitätsmedizin Berlin, Berlin, Germany | Heike Rössig |
| Institute of Tropical Medicine and International Health, Charité – Universitätsmedizin Berlin, Berlin, Germany | Maximilian Gertler |
| Charité Comprehensive Cancer Center, Charité – Universitätsmedizin Berlin, Berlin, Germany | Susen Burock |
| Department of Pediatric Surgery, Charité – Universitätsmedizin Berlin, Berlin, Germany | Katja von dem Busche |
| Berlin Institute for Clinical Teratology and Drug Risk Assessment in Pregnancy, Institute of Clinical Pharmacology and Toxicology, Charité – Universitätsmedizin Berlin, Berlin, Germany | Stephanie Patberg |
| Liverpool University Hospitals National Health Services Foundation Trust, UK & Department of Clinical Sciences, LSTM, UK | Angela Hyder-Wright |
| Department of Tropical Disease Biology, LSTM, UK | Alice J. Fraser |
| | Christopher T. Williams |
| | Edward I. Patterson |

| | |
|---|---|
| | Gala Garrod |
| | Grant A. Kay |
| | Grant L. Hughes |
| Department of Clinical Sciences, LSTM, UK | Elena Mitsi |
| | **Jesús Reiné** |
| | Ashleigh Howard |
| | Nikolaou Elissavet |
| | Carla Solorzano Gonzalez |
| | Lisa Hitchins |
| | Katerina Cheliotis |
| | Esther German |
| | Natalie Tate |
| | Sherin Pojar |
| Liverpool University Hospitals National Health Services Foundation Trust, UK & Department of Clinical Sciences, LSTM, UK | Helen Hill |
| Department of Clinical Sciences, LSTM, UK | Luis E. Cuevas |
| Department of Tropical Disease Biology, LSTM, UK | Rachel L. Byrne |
| | Sophie I. Owen |
| | Stefanie Menzies |

#### (B) Section: Questionnaire for study participants

**We invite you to participate in this survey. The survey serves to understand the diagnostic process and the disease and factors related to SARS-CoV-2 (novel coronavirus) infection.**

**Your answers will be kept strictly confidential and will not have a negative impact on your care. Participation in the study is voluntary and you have the option to skip questions that you do not want to answer.**

**The survey is expected to take 15-20 minutes. Thank you for your understanding and cooperation!**

| | |
|---|---|
| Postal code | *(Free text)* |
| Gender | ○ Male<br>○ Female<br>○ Diverse |
| How tall are you (in centimetres)? | *(Free text)* |
| How much do you weight (in kilograms)? | *(Free text)* |

**Symptoms that you attribute to the possible COVID-19**

| | |
|---|---|
| Did you have any symptoms of possible COVID-19 on the day of the test? | ○ No<br>○ Yes |
| Increased temperature / fever? | ○ No<br>○ Yes |
| Did you measure your fever? | ○ No<br>○ Yes |
| Highest temperature (in Celsius) | *(Free text)* |
| Cough | ○ No<br>○ Yes |
| Do you have a productive cough? | ○ No<br>○ Yes |
| Sore throat | ○ No<br>○ Yes |
| Shortness of breath | ○ No<br>○ Yes |
| Muscle pain / Body aches | ○ No<br>○ Yes |
| Fatigue | ○ No<br>○ Yes |

| | |
|---|---|
| Headache | ○ No<br>○ Yes |
| Runny nose | ○ No<br>○ Yes |
| Chest pain | ○ No<br>○ Yes |
| Diarrhea | ○ No<br>○ Yes |
| Nausea / vomiting | ○ No<br>○ Yes |
| Loss of taste or smell | ○ No<br>○ Yes |
| Other | ○ No<br>○ Yes |
| If yes, please specify | *(Free text)* |
| The earliest onset of symptoms attributed to possible COVID-19 | *(Day / Month / Year)* |
| How sick did you feel on the day of the test? | ○ Normal unrestricted activity as before the illness<br>○ Restriction with physical exertion, but able to walk; light physical work or work while sitting, e.g. light housework or office work, possible<br>○ Able to walk, self-sufficiency possible, but not able to work; can get up more than 50% of the waking time<br>○ Only limited self-sufficiency possible; 50% or more of the waking time tied to bed or chair<br>○ Completely in the need of care, no self-sufficiency possible; completely tied to bed or chair |
| Did you previously test negative? | ○ No<br>○ Yes |
| If yes, when | *(Day / Month / Year)* |
| If yes, where | ○ University clinic – inpatient<br>○ University clinic – outpatient<br>○ Drive-in<br>○ Other |
| Other, please specify | *(Free text)* |

| | |
|---|---|
| Do you know where you might have been infected with COVID-19? | ○ Household contact<br>○ Social contact<br>○ Work contact<br>○ Contact in university / school / kindergarten from you or a child in the family<br>○ Travel to risk area<br>○ Do not know<br>○ Other |
| Risk area | *(Free text)* |
| Other: please describe | *(Free text)* |

**Do you have any pre-existing conditions?**

| | |
|---|---|
| Which of the following lung disease(s) do you have? | ○ Asthma<br>○ Chronic Obstructive Pulmonary Disease – COPD<br>○ Breathing disorders during sleep<br>○ Obstructive Sleep Apnea – OSAS<br>○ Interstitial Lung Disease<br>○ Lung Cancer<br>○ Other<br>○ None |
| What other lung diseases do you have? | *(Free text)* |
| Cardiovascular diseases (e.g. hypertension, stroke, etc.) | ○ No<br>○ Yes |
| Chronic kidney disease | ○ No<br>○ Yes |
| Diabetes | ○ No<br>○ Yes |
| Autoimmune Disease (e.g. Rheumatoid Arthritis, MS) | ○ No<br>○ Yes |
| HIV | ○ No<br>○ Yes |
| Overweight | ○ No<br>○ Yes |
| Other, please specify | *(Free text)* |

#### (C) Section: Study Protocol

##### STUDY PROTOCOL

**Protocol Title**

Evaluation of the performance of novel rapid diagnostics for SARS-CoV-2 at point-of-care

**Short title**

Novel rapid diagnostics for COVID-19

**Protocol Version Number:**

Version 1.2

**Date:**

14th of April 2020

**Principal Investigator:**

Claudia Denkinger, MD
University Hospital Heidelberg
Division of Tropical Medicine
Im Neuenheimer Feld 324
69120 Heidelberg
Germany

**Co-Principal Investigator:**

Andreas Welker, MD

**Investigators:**

Britta Knorr

Lisa J. Krüger

Mary Gaeddert

Claudius Gottschalk

Frank Mockenhaupt

Andreas Lindner

**Financial support:** FIND – Provision of tests only

**Biometrician:** Lisa Köppel

**Conflict of interest**: None declared

**Confidentiality Statement:** The information contained in this document, especially unpublished data, is the property of University Hospital Heidelberg (or under its control) and may not be reproduced, published or disclosed to others without prior written authorization.

Institutions/Organizations/Partners Involved in the Study

**University Hospital Heidelberg (Principal Investigator)**
Claudia Denkinger, MD;
Division of Tropical Medicine
Im Neuenheimer Feld 324
69120 Heidelberg
Germany

**Gesundheitsamt (Co-Principal Investigator)**
Andreas Welker, MD
Gesundheitsamt des Rhein-Neckar-Kreises
Kurfürsten-Anlage 38-40
69115 Heidelberg

**Gesundheitsamt (Co-Investigator)**
Britta Knorr
Gesundheitsamt des Rhein-Neckar-Kreises
Kurfürsten-Anlage 38-40
69115 Heidelberg

**University Hospital Heidelberg (Co-Investigators)**
Lisa J. Krüger, Faculty of Medicine Mannheim
Mary Gaeddert, Division of Tropical Medicine
Claudius Gottschalk, Division of Tropical Medicine

**Charité – Universitätsmedizin Berlin (Co-Investigators)**
Frank Mockenhaupt and Andreas Lindner
Institute of Tropical Medicine and International Health
Campus Virchow-Klinikum
Augustenburger Platz 1
13353 Berlin

Statement of Principal Investigator

In signing this page, I, the undersigned, agree to conduct the study according to the principles outlined in the Declaration of Helsinki and in compliance with applicable regulatory requirements.

I will ensure that the requirements relating to obtaining Institutional Review Board (IRB)/ Independent Ethics Committee (IEC) review and approval are met. I will promptly report to the IRB/IEC any and all changes in the research activities covered by this protocol.

I have sufficient time to properly conduct and complete the study within the agreed study period and I have adequate resources (staff and facilities) for the foreseen duration of the study.

I am responsible for supervising any individual or party to whom I delegate study related duties and functions conducted at the study site. Further, I will ensure this individual or party is qualified to perform those study-related duties and functions.

I understand that all information obtained during the conduct of the study with regard to the subjects' state of health will be regarded as confidential. No participant's names or personal identifying information may be disclosed. All participant data will be anonymized and identified by assigned numbers on all documents. Monitoring and auditing and inspection by the appropriate regulatory authority(ies), will be permitted.

I will maintain confidentiality of this protocol and all other related investigational materials. Information taken from the study protocol may not be disseminated or discussed with a third party.

Name of Principal Investigator:

Signature: ________________________________ Date: ________________________

DD/MMM/YYYY

#### Protocol History/Amendment Summary

| Version number | Release date | Comments |
|---|---|---|
| 1.0 | 21.03.2020 | Initial version |
| 1.1. | 24.03.2020 | Recommendations from ethic committee |
| 1.2. | 14.04.2020 | Inclusion of a new study site in Berlin |

### List of Abbreviations and Acronyms

| **Abbreviation/acronym** | **Meaning** |
|---|---|
| COVID-19 | Corona-Virus-Disease-19 |
| FIND | Foundation for Innovative New Diagnostics |
| POC | Point-of-care |
| RT-PCR | Real-Time Polymerase-Chain-Reaction |
| SARS-CoV2 | Severe acute respiratory syndrome coronavirus *2* |
| PPE | Personal Protection Equipment |
| PI | Principal Investigator |
| NP swab | Nasopharyngeal swab |
| OP swab | Oropharyngeal swab |

#### Protocol Synopsis

| | |
|---|---|
| **Title** | Evaluation of the performance of novel rapid diagnostics for SARS-CoV-2 at point-of-care |
| **Rationale and background** | The aim of this study is to validate novel rapid point-of-care (POC) tests that can determine if a person has COVID-19, a serious and sometimes fatal respiratory infection caused by the coronavirus SARS-CoV-2.<br>The COVID-19 outbreak has rapidly spread across the globe including Germany. As of March 18th 2020, there are a total of 8.198 cases all diagnosed by high-complexity nucleic acid amplification testing (RT-PCR) in Germany. However, the consensus among public health officials is that the number of infected individuals is far higher. Given the wide range of possible symptoms and the potential for transmission before individuals are aware that they are infected, exposure to SARS-CoV-2 is a particular hazard for health care providers. Lack of diagnostic testing capacity and the long turnaround time for return of results of the current gold standard for testing (RT-PCR) have interfered with the ability of public health officials to track and contain the disease. The lack of capacity, in turn, is due in part to the logistic challenges and global reagent shortages faced by laboratories attempting to implement new RT-PCR assays for SARS-CoV-2.<br>Rapid tests for SARS-CoV-2, if shown to have sufficient accuracy to aid in clinical decision-making, could contribute substantially to control of disease spread globally. In particular, rapid tests might provide the only source of diagnostic testing in low-and middle-income countries unable to implement RT-PCR.<br>The first novel POC tests to be validated in this study are manufactured by Bioeasy (Guangdong Province, China) and SD BIOSENSOR (Suwon, South Korea). Other tests will be considered as they become available based on analytical testing in partner laboratories with simulated and/or banked samples prior to selection for clinical validation. Novel rapid tests will not be used for clinical decision making. |
| **Use case of test** | Accuracy of diagnostic testing in adults with suspected COVID-19 infection |
| **Primary objective** | *To determine the diagnostic accuracy of COVID-19 antigen tests on a respiratory specimen (NP swab, OP swab), vs gold-standard real-time reverse-transcription PCR (RT-PCR), as performed in an affiliated reference laboratory on a respiratory specimen.* |
| **Secondary objective** | To assess the feasibility, ease of use of the index test at POC in comparison to in laboratory use |
| **Exploratory objective** | *To determine the diagnostic accuracy of COVID-19 antigen tests on a respiratory specimen (buccal swab and saliva), vs gold-standard real-time reverse-transcription PCR (RT-PCR), as performed in an affiliated reference laboratory on a respiratory specimen compared with the index on NP and OP samples*<br>*To determine the association of positive index test results with mortality* |
| **Study design & Participants** | This is a prospective study for diagnostic accuracy. The different test will be evaluated in substudies. The same study design will be applied to each test evaluation (substudy) performed under this protocol. Each substudy is a separate diagnostic accuracy study, with the primary objective of validating the performance of a novel rapid POC SARS-CoV-2 test in patients with suspected COVID-19 presenting to be tested at the testing sites.<br>We will enroll at least 500 participants and a maximum of 2,000 participants suspected to have COVID-19 at all sites per each substudy. At least 2,000 participants with a maximum of 5,000 participants in total.<br>Participants included will meet testing criteria as determined by the Department of Public Health. |
| **Location** | Established testing sites of the Department of Public Health in Heidelberg and surroundings.<br>A further study site is planned at Charité – Universitätsmedizin Berlin |

| | |
|---|---|
| **Duration** | March 2020 – March 2021 |

#### Protocol

#### Rationale and Background

The aim of this study is to validate novel rapid POC tests that can determine if a person has COVID-19, a serious and sometimes fatal respiratory infection caused by the coronavirus SARS-CoV-2. Persons with COVID-19 may develop fever, cough, and shortness of breath, in addition to other symptoms (sore throat, diarrhea). Case severity ranges from mild to fatal.

The COVID-19 outbreak has rapidly spread across the globe including Germany, with cases of COVID-19 confirmed in all sixteen federal states. As of March, 21st 2020, there are a total of 16.662 cases all diagnosed by high-complexity nucleic acid amplification testing (RT-PCR) in Germany. However, the consensus among public health officials is that the number of infected individuals is far higher. Given the wide range of possible symptoms and the potential for transmission before individuals are aware, they are infected, exposure to SARS-CoV-2 is a particular hazard for health care providers. Lack of diagnostic testing capacity and the long turnaround time for return of results have interfered with the ability of health care providers to conserve personal protective equipment (PPE) and the ability of public health officials to track and contain the disease. The lack of capacity, in turn, is due in part to the logistic challenges and global reagent shortages faced by laboratories attempting to implement new RT-PCR assays for SARS-CoV-2. Rapid tests for SARS-CoV-2, if shown to have sufficient accuracy to aid in clinical decision-making, could contribute substantially to control of disease spread in Germany and globally. In particular, rapid tests might provide the only source of diagnostic testing in low-and middle-income countries unable to implement RT-PCR.

In the absence of a vaccine or effective treatment, prevention remains the mainstay of epidemic control. This includes early isolation of infectious cases, in turn requiring accurate diagnosis. The current need for specialized laboratories for RT-PCR testing, with people aggregating in front of testing sites, creates barriers to testing, making symptomatic individuals less likely to present for testing and accelerating community transmission.

In this context, SARS-CoV-2 testing using rapid POC tests holds great potential. A test that can reliably detect early infection at POC can facilitate rapid case identification and isolation, reduce the risks of transmission, and make decentralized SARS-CoV-2 testing data more readily available to inform prevention measures.

Several novel POC tests have been developed. With assistance from the Foundation for Innovative New Diagnostics (FIND), we will identify several candidate tests for validation. Each test will be evaluated in a parallel sub-study, and compared to the gold standard of RT-PCR.

The clinical hypothesis is that the novel rapid POC test evaluated in each substudy will maintain clinical performance of $\geq$90% sensitivity and $\geq$95% specificity when compared to RT-PCR performed on a respiratory sample in the central (reference) laboratory. Novel rapid tests will not be used for clinical decision making.

The study presented will evaluate several promising tests in parallel.

#### Study Objectives and Endpoints

The purpose of this study is to evaluate the performance of newly-developed rapid diagnostic tests for POC diagnosis of infection with SARS-CoV-2, a novel coronavirus responsible for the disease known as COVID-19. Specifically, we will compare the performance of novel tests [for detection of SARS-CoV-2 nucleic acid or antigen], presumably suitable for POC testing to the current gold standard testing [RT-PCR, as performed in a reference laboratory] in suspected SARS-CoV-2 cases presenting to testing sites and meeting criteria for testing. We will evaluate each novel POC test in a separate, dedicated evaluation (substudy) to facilitate rapid evaluation of test performance; each substudy will have the same design, as outlined below. Results of novel POC tests will not be used for clinical management. For each novel POC test evaluated, we will investigate the following objectives:

**Primary Objective:** Validate sensitivity and specificity of the novel POC SARS-CoV-2 tests, as performed on a respiratory specimen (nasopharyngeal swab, oropharyngeal swab vs. gold-standard real-time reverse-transcription PCR (RT-PCR)), as performed in an affiliated reference laboratory on a respiratory specimen. The novel POC SARS-CoV-2 test may also be performed in the reference laboratory as a control.

**Secondary Objective:** Evaluate feasibility and ease-of-use of the novel POC SARS-CoV-2 test at POC.

**Exploratory Objective:** Validate sensitivity and specificity of the novel POC SARS-CoV-2 test, as performed at POC on a saliva/buccal swab sample, vs gold-standard RT-PCR, as performed in an affiliated reference laboratory on a respiratory sample.

**Hypothesis:** Each novel SARS-CoV-2 rapid diagnostic test, when performed at POC on a respiratory sample, will maintain clinical performance of $\geq$90% sensitivity and $\geq$95% specificity when compared to RT-PCR performed at an affiliated reference laboratory.

**Table 1. Study Objectives and Endpoints**

| OBJECTIVES | ENDPOINTS |
|---|---|
| **PRIMARY** | |
| 1.1. *To determine the diagnostic accuracy of COVID-19 antigen tests on a respiratory specimen (NP swab, OP swab), vs gold-standard real-time reverse-transcription PCR (RT-PCR), as performed in an affiliated reference laboratory on a respiratory specimen.* | 1.1. Point estimates of sensitivity and specificity of index test, with 95% confidence intervals, using an RT-PCR reference standard |
| **SECONDARY** | |
| 2.1 To assess the feasibility, ease of use of the index test at POC in comparison to in laboratory use | 2.1. Time to proficiency, implementation issues, design related issues at POC |
| **EXPLORATORY** | |
| 3.1. *To determine the diagnostic accuracy of COVID-19 antigen tests on a respiratory specimen (buccal swab and saliva), vs gold-standard real-time reverse-transcription PCR (RT-PCR), as performed in an affiliated reference laboratory on a respiratory specimen compared with the index on NP and OP samples*<br>3.2. *To determine the association of positive index test results with mortality* | 3.1. Point estimates of sensitivity and specificity of index test, with 95% confidence intervals, using an RT-PCR reference standard<br>3.2. *Survival analysis for the outcome of death within 2-3 months by COVID and Antigen test status* |

#### Study design

This is a prospective study for diagnostic accuracy. The same study design will be applied to each test evaluation (substudy) performed under this protocol. Each substudy is a separate diagnostic accuracy study, with the primary objective of validating the performance of a novel rapid POC SARS-CoV-2 test in patients with suspected COVID-19 presenting to be tested at testing sites.

##### 3.1 Subject Population

We will enroll at least 500 participants per each substudy and a maximum of 2,000 participants suspected to have COVID-19 at all sites. At least 2,000 participants with a maximum of 5,000 participants in total. Interim analyses are planned to decide on continuation of testing using predefined criteria. See sample size and sample analysis discussed Section 8.

##### 3.2 Inclusion Criteria

Any person ≥ 18 years of age presenting to temporary testing locations around Heidelberg, Germany who has voluntarily given written consent and is willing to participate in this study. The Department of Public Health has defined criteria for who should be tested at these testing sites as part of routine medical care. The current criteria (as of 20th of March 2020) is to only test individuals who have been in direct contact with confirmed COVID-19 cases or individuals coming from a high-risk area. These criteria from the Department of Public Health might be subject to changes in the future. The population presenting at the test site meets the Department of Public Health definition of suspected COVID-19 cases and is thus being tested for SARS-CoV-2 as part of routine medical care.

##### 3.3 Exclusion Criteria

- Hemodynamic instability as determined by the treating physician
- Patient unable to cooperate with respiratory sample collection
- Patient unable to give informed consent
- Patient receiving a confirmatory second test after a first positive diagnosis for SARS-CoV-2
- Recent history of excessive nose bleeds

##### 3.4 Study location

Established testing sites of the Department of Public Health in Heidelberg and surroundings.

Furthermore, a study site is planned at the testing site of Charité – Universitätsmedizin Berlin.

If study participants are insufficiently recruited, further study sites may be considered in future.

### 4 Study intervention

#### 4.1 Investigational product

The first novel POC tests to be studied under this protocol are manufactured by Bioeasy (Guangdong Province, China) and SD BIOSENSOR (Suwon, South Korea) [see Appendix]. Other tests will be considered as they become available based on analytical testing in partner laboratories with simulated and/or banked samples prior to selection for clinical validation under this protocol and will have been vetted through extensive discussions with the Foundation of Innovative New Diagnostics (FIND), the WHO Collaborating Center for COVID diagnostic test evaluation.

#### 4.2 Comparator product

Comparator product will be the current standard of care RT-PCR in each setting. At the University Hospital Heidelberg, this is currently performed on a QiaSymphony for automated extraction and TipMolbiol on a Roche Lightcycler for RT-PCR. Other comparable RT-PCRs might be implemented throughout the study but will be considered equivalent in respect to the study. If possible, all subjects enrolled for testing with a given novel POC test will have their RT-PCR testing done with the same RT-PCR protocol in the same laboratory.

#### 4.3 Preparation/Handling/Storage/Accountability

##### 4.3.1. Acquisition

Procurement of the investigational products will be done through FIND, who will coordinate shipments from the manufacturer. The study site will maintain an updated inventory of the trial materials and will inform FIND immediately if additional materials are required.

The investigator or designee must confirm that appropriate temperature conditions have been maintained during transit for the investigational product received and any discrepancies will be reported and resolved before its use.

##### 4.3.2. Storage

The investigational product will be stored in a secure, environmentally controlled, and monitored (manual or automated) area in accordance with the labelled storage conditions with access limited to the investigator and authorized site staff.

##### 4.3.3. Test Handling and Performance

Testing using the investigational products will be performed according to the manufacturer’s instructions.

Only NP or OP samples from participants enrolled in the trial will be processed with the investigational products for the main aims. For exploratory aims, buccal swab and saliva will be considered for testing as it would enable self-testing but sensitivity would be expected to be lower.

##### 4.3.4. Accountability

The investigator is responsible for trial intervention accountability, reconciliation, and record maintenance (i.e., receipt, reconciliation, and final disposition records). Investigational Product Accountability logs filled at each site will ensure the proper follow-up of the used, failed and remaining investigational products

##### 4.3.5. Export and Import Permits

The study investigator is responsible for making import permit applications in a timely manner. FIND logistics team will support.

###### 4.3.6. Quality Control Check for Incoming Shipments

Upon arrival of each new shipment of assays, the sites will conduct and document an incoming quality check following a separate SOP. New lots may only be used after this quality check is successfully passed.

#### 4.4 Minimization of Error and Bias

##### 4.4.1 Patient selection

Spectrum bias will be avoided by enrolling a consecutive series of study participants, and by using a prospective trial design. Enrolment will be based on clearly defined eligibility criteria. If most testing is performed in ambulatory testing sites, the most severely ill patients cannot be enrolled, as this patient group will not present itself there. This may lead to a degree of underestimation of sensitivity. However, to ensure the validity and generalizability of study results, descriptive statistics on patient characteristics and estimates of diagnostic accuracy will be reported separately for relevant subgroups (outpatients/inpatients, duration of symptoms and severity of symptoms) that are a proxy for disease spectrum.

##### 4.4.2 Index test

The overall risk of review bias is minimal even if the index test result requires subjective interpretation (e.g. band intensity) as the personnel interpreting recording the results will be blinded to all other test results. Harmonization of result interpretation will be ensured with proper training, proficiency assessment and competency assessment at the beginning of the study.

##### 4.4.3 Reference standard

The reference standard RT-PCR is considered to have near perfect sensitivity and specificity. Thus, a bias is unlikely.

##### 4.4.4 Flow and timing

Samples for index test testing will be collected in parallel to the samples that will be used for reference testing so disease progression bias is not a concern.

##### 4.4.5 Independence of the investigators

All aspects of the study, including specimen collection, testing, data entry, and data analysis, will be performed independently of the manufacturer of the novel POC test under study. All de-identified clinical and laboratory data will be analyzed by Dr. Claudia Denkinger's study team, which has no financial ties/commercial interests or personal conflicts of interest related to any participating test manufacturers.

### 5 Study procedures

#### 5.1 Specimen collection and location of testing

A respiratory sample will be collected on the same day by NP swab, OP swab, or buccal swab for POC testing, as appropriate to the novel POC test under evaluation using personal protection equipment in a containment zone as recommend by the University Hospital Heidelberg SOP with strict adherence to the manufacturer's SOP. A respiratory sample will be collected in parallel on the same day for routine testing at the central laboratory at University Hospital Heidelberg per clinical routine. Written consent will be obtained from all subjects whose samples will be used for POC testing.

Enrolled patients receive a study number to reidentify their laboratory specimens and clinical data.

The clinical respiratory sample (collected in parallel with samples for POC testing) will be transported and tested to the central laboratory using RT-PCR, per standard practice (below).

Patients will be diagnosed based on RT-PCR results and clinical signs and symptoms by their treating physician; *results of novel POC tests will not be used for clinical management*.

Processes for respiratory sample collection for SARS-CoV-2 diagnostic testing are routine and well-practiced in Germany. Health care workers collecting samples are already trained specifically in respiratory sample collection techniques and are wearing appropriate personal protective equipment (PPE) at the time of sample collection. All novel POC tests will be performed within the already designated isolation zones for patients suspected of having SARS-CoV-2, meaning that everyone in that area will similarly already be wearing appropriate PPE per clinical routine, and the testing will not add any new biosafety hazard to routine clinical practices.

#### 5.2 Data collection

To support validation of assay performance, we will collect demographic and clinical data for each participant via telephone while the participant is in a safe home surrounding. A questionnaire will collect data on age, sex, symptoms, number of days symptomatic at the time of testing, and severity of illness.

Laboratory results of the novel POC tests, including positive/negative, valid vs invalid, reason for invalid, band density (for lateral flow tests), will be recorded on-site. Also, the ambient temperature and humidity in the testing location will be recorded.

Results of the routine RT-PCR tests, including Ct values, will also be obtained from the University Hospital Heidelberg electronic medical record and accordingly at new study sites, like Charité – Universitätsmedizin Berlin. Results of other virologic tests like influenza will be recorded if available. Patient outcomes, including mortality, will be recorded from hospital or public health records.

We will also collect operator feedback on the ease of test operation and results interpretation. All feedback will be logged via a System Usability Survey (SUS) scoring system.

#### 5.3. Data Management

Electronic data management from the University of Heidelberg and RedCap and accordingly at new study sites, like Charité – Universitätsmedizin Berlin.

Data management: Clinical data and results of the novel diagnostic tests will be recorded on paper forms and entered into an electronic database (RedCap). Other information on the novel diagnostic tests, including user proficiency testing and storage conditions, will be recorded systematically. Standard Operating Procedures (SOPs) will be developed for screening, sample collection, and laboratory testing.

### 6 Participant Discontinuation/Withdrawal

A participant may be withdrawn at any time at the discretion of the investigator for safety, behavioural, compliance, or administrative reasons.

If the participant withdraws consent for disclosure of future information, FIND may retain and continue to use any data collected before such a withdrawal of consent in the case that the participant voluntarily agrees to the usage of the prior to the withdrawal of consent collected data. The participant may request the deletion of all prior collected data and the destruction of any samples. In this case the investigator must document this in the site trial records

If a participant withdraws from the trial, he/she may request destruction of any samples taken and not tested, and the investigator must document this in the site trial records.

### 7 Safety and Incident Reporting

Given that this is a diagnostic accuracy study that is not utilizing test results for patient care in clinical decision making and given that additional procedures for the trial, i.e. NP/OP collection are extremely low risk, the probability of an AE or SAE occurring to a trial participant to be associated with the investigational products is extremely low.

This study does not have predefined termination criteria being an accuracy study.

#### 7.1. Adverse Events and Serious Adverse Events

The definitions of an Adverse Event (AE) and Serious Adverse Event (SAE) are considered as per MEDDEV 2.7/1 Rev 4.

Given the nature of this trial AE reporting is limited in scope to:

- SAEs that may be associated with NP/OP collection.
- SAEs that occur at the testing sites using the investigational product (see section 7.4 Medical device incidents).
- Any other serious events that affect the rights safety or welfare of subjects.

#### 7.2. Time Period for Collecting SAE Information

Information will be collected at the specimen collection and in the testing of the investigational product. SAEs will be recorded and reported to the sponsor or designee within 24 hours of the occurrence. The investigator will submit any updated SAE data to the sponsor within 24 hours of being made aware of the event.

#### 7.3. Reporting and Follow up of SAEs

The PI has a legal responsibility to notify both the local regulatory authority and other regulatory agencies about the safety of a trial intervention under clinical investigation. The PI will comply with country-specific regulatory requirements relating to safety reporting to the regulatory authority, Institutional Review Boards (IRB)/Independent Ethics Committees (IEC), and investigators.

An investigator who receives a safety report describing a SAE or other specific safety information (e.g., summary or listing of SAEs) from FIND will review and then file it in the Investigator Site File (ISF) and will notify the IRB/IEC, if appropriate according to local requirements.

##### 7.4. Medical Device Incidents (including Malfunctions)

Medical devices are being provided for use in this study. In order to fulfil regulatory reporting obligations worldwide, the investigator is responsible for the detection and documentation of events meeting the definitions of incident or malfunction that occur during the trial with such devices.

Given that most POC tests will be instrument free. This is of limited concern for this study.

### 8 Study Analysis

The statistical analysis plan (SAP) will be developed and finalized before the start of enrolment; this section details a summary of the planned statistical analyses of the primary and secondary endpoints, as well as the rationale for the proposed sample size. The statistical analysis will be conducted with STATA, a software for statistical analysis and data science.

#### 8.1 Populations for Analysis

For purposes of analysis, the following populations are defined:

Table 3: Populations for Analysis

| Population | Description |
|---|---|
| Enrolled/Intention-to-test (ITT) | All subjects successfully enrolled in the study (having signed the ICF) |
| Evaluable/Per Protocol Population (PP) | All subjects in ITT who have samples available, and valid results for all tests |
| Survival population | For subjects in ITT who have data on vital status at 2-3 month from death register |

#### 8.2 Statistical Analyses

General Methodology: Point estimates of sensitivity and specificity, with 95% confidence intervals based on Wilson’s score methods, will be calculated following the definitions:

Table 4: Case predictions

| | Reference standard classification | | | |
|---|---|---|---|---|
| Case prediction | | Positive | Negative | Total |
| | Predicted positive | a | b | (a + b) |
| | Predicted negative | c | d | (c + d) |
| | Total | (a + c) | (b + d) | (a + b + c + d) |

Table 5: Sensitivity and Specificity

| | |
|---|---|
| **a = True Positives,**<br>**b = False Positives**<br>**c = False Negatives**<br>**d = True Negatives** | Sensitivity = a / (a + c)<br><br>Specificity = d / (b + d) |

Endpoints: See table with objectives and endpoints above

Subgroup analysis:

- By duration of symptoms (as a categorical variable)
- By severity of symptoms (as a categorical variable)
- Outpatient vs. inpatients
- Age (as a categorical variable)

The endpoints on accuracy will be calculated on the PP.

Kaplan–Meier survival curves will be generated to investigate the risk of mortality at 2-3 months, based on index test results among all patients where vital status assessment can be done.

Feasibility will be assessed by asking the POC heath care worker to fill a questionnaire about design, ease of use and implementation of the index test

Further detail of an analysis will be described in the SAP.

##### 8.3 Interim Analysis

We have chosen sample sizes such that good tests with sensitivity and specificity at or above minimum values of interest have a high likelihood of being advanced (i.e., high power), and poor tests with sensitivity and specificity at or below maximum values of NO interest have a low likelihood of being advanced (i.e. low false-positive rate).
An interim analysis will be performed after 25%, 50% enrollment and decisions will be made on advancement of the test for further testing based on calculated sensitivity and specificity. If a test does not reach at least 90% sensitivity (point estimate), the added value will be considered in terms of time to diagnose and ease of implementation. Tests with sensitivity below 70% (point estimate) will not be evaluated further.

##### 8.4 Sample Size Determination

The target sample size was chosen to achieve an acceptable level of precision for the estimates of index test sensitivity and specificity. It has been assumed that the average expected sensitivity for index test should be 95%, based on data previously gathered by the manufacturer. The average expected overall prevalence ranges between 1 to 10%. Based on these considerations, a sample size of 2000 patients would yield sensitivity estimates with a precision of +/- =10% at prevalence of 1%, precision of +/- =4% at prevalence of 5% and precision of +/- =3% at prevalence of 10% at a significance level of alpha = 0.05 (corresponding to 95% confidence interval). Given the large number of test-negative participants, the estimate of specificity will be precise.

#### 9 Benefits

There will be no direct benefit to subjects from this research. All study personnel will be made aware that the novel POC tests under study are for research purposes only and cannot be used to determine whether or not to initiate treatment, nor for any other clinical management decisions. As discussed above, development of COVID-19 POC tests will improve diagnosis and treatment of COVID-19, facilitate studies to understand its prevalence and natural history, and ultimately lead to effective vaccines and therapies.

#### 10 Remuneration

There will be no payment for participation in this research study.

#### 11 Costs

There will be no costs to the subject for participating in this research study.

#### 12 Alternatives

The alternative is that the subjects do not have to participate in the research.

#### 13 Ethical Considerations

##### 13.1. Ethics Approval

This study will be conducted in accordance with the protocol and with the following:

- Consensus ethical principles derived from international guidelines including the Declaration of Helsinki (seventh revision, version 2013)
- Applicable laws and regulations
- Before the start of the study, the investigators will provide the following documents:
- Curriculum vitae of the Principal Investigator
- Protocol Signature Page, signed and dated by the PI
- IRB/IEC approval letter from the Medical Faculty Heidelberg and the local IRB/IEC for the study protocol and written consent form.

At the extended study site in Berlin, the local ethics committee of the Charité - Universitätsmedizin Berlin will be involved, as well as the Charité data protection officer (behördlicher Datenschutzbeauftragter). Further sites in the future would accordingly involve the local ethics committees and data protection officers.

##### 13.2. Informed Consent Process

Eligible potential subjects will be recruited and witnessed, documented written informed consent will be obtained by local clinical personnel/study staff trained in human subject's protections. Recruitment will occur at the time that the patient is approached for OP/NP swab collection for testing per clinical routine. Strict infection control measures are in place at all testing centers. These measures require that all items coming in contact with suspected patients will be handled in full PPE and subsequently considered infectious waste. A written consent script describing confidentiality regulations and a participant information sheet describing the study purpose and procedures will be given to each subject. The written consent form will be signed by the participant and returned to the study personnel at the testing site. Identified as infectious paper, this written consent form will be documented and digitally saved in a password- protected data management system and disposed at the site.

##### 13.3. Subject confidentiality

Careful attention will be paid to maintaining confidentiality of all paper documents, electronic data, and biological specimens. Participants will each be assigned a unique ID code, which will be linked to their name on a password-protected form stored on a password-protected study computer. This code will be used in lieu of participants' names on all hard copy forms, electronic datasets and biological specimens.

##### 13.4. Data Protection

Data will be stored in a secure way accessible only to the researchers involved: password encrypted (digital). In case of incidental findings related to patients diagnoses, participant will be referred to a person able to correct the problem or clarify the matter (i.e. counsellor, nurse). All the information collected for this study will be kept strictly confidential by identifying the participants with a unique code (or study ID) to which only the study investigators have access. Only de-identified data will be kept on a password protected server for at least 5 years following Proposals for Safeguarding Good Scientific Practice (version 2013) of the German Research Foundation (or longer if local regulations require). Names of participants and other personal data is subject to data protection according to the General Data Protection Regulation (GDPR) as well as local and German federal and state data protections laws. Information may only be passed on to other institutions involved in the study if they are pseudonymized for data protection reasons. All data generated in this study, absent any personally identifiable information, may be sent to outside collaborators for further analysis. Personal information and names will not be revealed at publication and all personal data will be anonymized accordingly to the research purpose.

##### 13.5. Publication Policy

Data obtained from participation in this study are considered confidential. The investigators must adhere to the non-disclosure requirements set forth in the contractual agreement.
Participants of the study have the opportunity to be informed about the general outcome of the study. To do so, they are asked to contact an investigator. Furthermore, the study will be registered on German Clinical Trials Register.
(www.drks.de).

Authorship for scientific publication of the study results will be determined by mutual agreement and in line with International Committee of Medical Journal Editors authorship requirements, as described in the publication policy section of the contractual agreement.

#### Appendices

**Schedule of Activities**

| **Milestones** | **Timeline** (weeks) | | | | | | | | | | | |
|---|---|---|---|---|---|---|---|---|---|---|---|---|
| **Activities** | 1 | 2 | 3 | 4 | 5 | 6 | 7 | 8 | 9 | 10 | 11 | 12 |
| Research Preparation<br>○ Establish flow at testing site<br>○ Obtain materials<br>○ Design data tools and database<br>○ Ethics approval obtained | x | x | | | | | | | | | | |
| Participant Enrollment<br>Sample collection and testing at field site | | | x | x | x | | x | x | x | | | |
| Primary Data Analyzed<br>Final Data Analysis | | | | | x | x | x | | | x | x | x |
| Knowledge Disseminated and Translated<br>Draft report written<br>Report submitted to partners and WHO<br>Draft Manuscripts written<br>Manuscripts submitted | | | | | | | x | x<br>x | x | x | | |

### (D) Section: Ease of Use Assessment Questionnaire

#### Usability evaluation - part II

"Evaluation of the performance of novel rapid diagnostics for SARS-CoV-2 at point-of-care"

Thank you for your time to answer this questionnaire (about 20 minutes).

*Your input is very valuable!*

***OVERALL QUESTIONS***

1. **User identifier**

2. **Date of filling the questionnaire**

3. **In which country do you currently work?**

4. **At which facility / study site do you currently work?** *Mark only one oval.*

   - Schwetzingen (Heidelberg)
   - Berlin
   - Liverpool

5. **Which test are you assessing?** *Mark only one oval.*

   - Coris Bioconcept Respi Strip
   - Bioeasy FIA
   - Bioeasy Colloidal Gold
   - SD Biosensor Standard F (Flourescence)
   - SD Biosensor Standard Q
   - Rapigen Biocredit Colloidal Gold

6. **About how many times did you perform this test approximately?** *Mark only one oval.*

- Only observed use
- < 10
- 10 – 100
- > 100

7. **About how many times did you observe the use of this test (not performed yourself)?** *Mark only one oval.*

- < 10
- 10 – 50
- 50 – 100
- > 100

8. **What is your profession?**

9. **How many years of laboratory experience do you have?**

10. **How many years of working experience in limited resource settings do you have?**

11. **How much experience do you have with interpreting the results of lateral flow tests or rapid diagnostics (e.g. for HIV, malaria, pregnancy)?** Please note that we refer here to your experience with INTERPRETING the test results. If you do not conduct the test yourself, but do inform patients about the test results, we also consider that as experience with INTERPRETING the test results. *Mark only one oval.*

- None
- < 1 year
- 1 – 3 years
- > 3 years

***TEST SPECIFIC QUESTIONS***

*TRAINING*

**12. How satisfied were you with the following components of the test training?** *Mark only one oval per row.*

| | Very satisfied | Satisfied | Neither | Dissatisfied | Very dissatisfied |
|---|---|---|---|---|---|
| Instructions for Use | ⬭ | ⬭ | ⬭ | ⬭ | ⬭ |
| Standard Operating Procedures | ⬭ | ⬭ | ⬭ | ⬭ | ⬭ |
| Face to face demonstration | ⬭ | ⬭ | ⬭ | ⬭ | ⬭ |

**13. What additional materials (if any) do you think should be provided as part of the training?**

- None
- Other:

**14. How long should be the training of this test?** *Mark only one oval.*

- Self-explanatory, no need for training
- 1 - 2 hours
- 2 - 4 hours
- Half a day
- Full day

**15. Do you consider proficiency testing necessary?**

Proficiency testing as in assessing the user's performance or ability to run the test following the training. *Mark only one oval.*

- Yes
- No
- Other: :

**16. Please comment here on the need for proficiency testing**

**17. After how many of these tests do you feel you could perform the test on your own (having access to the training material)?** *Mark only one oval.*

- ⬭ 1 – 2 tests
- ⬭ 3 – 5 tests
- ⬭ 6 – 10 tests
- ⬭ > 10 tests

*ASSESSMENT OF TEST*

**18. How satisfied are you with the quality of each of the components in the kit (in terms of ease of use and fit for purpose)?** *Mark only one oval per row*

| | Very satisfied | Satisfied | Neither | Dissatisfied | Very dissatisfied |
|---|---|---|---|---|---|
| External paper box of kit | ⬭ | ⬭ | ⬭ | ⬭ | ⬭ |
| Assay diluent tube | ⬭ | ⬭ | ⬭ | ⬭ | ⬭ |
| Filter cap (if necessary) | ⬭ | ⬭ | ⬭ | ⬭ | ⬭ |
| Swab for specimen collection | ⬭ | ⬭ | ⬭ | ⬭ | ⬭ |
| Test cartridge / device | ⬭ | ⬭ | ⬭ | ⬭ | ⬭ |
| Test cartridge packing / pouch | ⬭ | ⬭ | ⬭ | ⬭ | ⬭ |
| Reader (if necessary) | ⬭ | ⬭ | ⬭ | ⬭ | ⬭ |

**19. Which kit component(s) should be improved in your opinion (if any)?** *Please specify why and how*

**20. Overall, how satisfied are you with the kit components?** *Mark only one oval.*

| | 1 | 2 | 3 | 4 | 5 | |
|---|---|---|---|---|---|---|
| Very satisfied | ⬭ | ⬭ | ⬭ | ⬭ | ⬭ | Very dissatisfied |

**21. How satisfied are you with the overall design of the device in terms of the following features?** *Mark only one oval per row.*

| | Very satisfied | Satisfied | Neither | Dissatisfied | Very dissatisfied |
|---|---|---|---|---|---|
| Size of cartridge | ⬭ | ⬭ | ⬭ | ⬭ | ⬭ |
| Space for labeling on the front | ⬭ | ⬭ | ⬭ | ⬭ | ⬭ |
| Size of the well to add sample mix | ⬭ | ⬭ | ⬭ | ⬭ | ⬭ |
| Size of reading window | ⬭ | ⬭ | ⬭ | ⬭ | ⬭ |
| Logical sequence of steps | ⬭ | ⬭ | ⬭ | ⬭ | ⬭ |

**22. Overall, how satisfied are you with the time relevant components?** *Mark only one oval.*

| | 1 | 2 | 3 | 4 | 5 | |
|---|---|---|---|---|---|---|
| Very satisfied | | | | | | Very dissatisfied |

**23. Please assess the Test's storage conditions.** *Mark only one oval.*

| | > 12 months | 12 to 6 months | 5 to 3 months | < 3 months |
|---|---|---|---|---|
| Stability of test | | | | |
| Stability of control material (if appl.) | | | | |

| | 2 – 40° | 15 – 35° | 15 – 30° | 20 – 25° |
|---|---|---|---|---|
| Storage temperature | | | | |

**24. Overall, how satisfied are you with the test's storage conditions?** *Mark only one oval.*

| | 1 | 2 | 3 | 4 | 5 | |
|---|---|---|---|---|---|---|
| Very satisfied | | | | | | Very dissatisfied |

**25. Which part(s) of the device could be improved in your opinion (if any)?**
*Please specify why and how.*

**26. Please determine the difficulty of the following steps:**

Please consider your day-to-day/routine workload (or that of the people in the lab/area where this test could be implemented) to answer this question.

*Mark only one oval per row.*

| | Very easy | Easy | Neither | Difficult | Very difficult |
|---|---|---|---|---|---|
| a) Check expiry date | | | | | |
| b) Remove the test cartridge from the pouch | | | | | |
| c) Label the test cartridge with patient identifier | | | | | |
| d) Label the assay diluent tube with patient identifier | | | | | |
| e) Open the assay diluent tube by removing the seal (if appl.) | | | | | |
| f) Transfer of buffer into diluent tube (if applicable) | | | | | |
| g) Insert the swab into the tube | | | | | |
| h) Ease of swab extraction procedure | | | | | |
| i) Ability to perform swab extraction procedure consistently | | | | | |
| j) Ability to maintain cleanliness of ancillary devices (e.g. pipette) in order to avoid cross contamination | | | | | |
| k) Ease of transferring sample onto device | | | | | |
| l) Ease of transferring exact quantity into the sample well | | | | | |
| m) Trouble shooting | | | | | |

**27. How satisfied are you with the logical sequence of steps?** *Mark only one oval.*

| | 1 | 2 | 3 | 4 | 5 | |
|---|---|---|---|---|---|---|
| Very satisfied | | | | | | Very dissatisfied |

**28. Overall, how difficult did you find the steps?** *Mark only one oval.*

| | 1 | 2 | 3 | 4 | 5 | |
|---|---|---|---|---|---|---|
| Very easy | ◯ | ◯ | ◯ | ◯ | ◯ | Very difficult |

**29. Please assess the time relevant components of the test.** *Mark only one oval per row.*

| | ≤ 2 min | 3 to 5 min | 6 to 10 min | > 10 min |
|---|---|---|---|---|
| Pre analytic time | ◯ | ◯ | ◯ | ◯ |
| Analytic time | ◯ | ◯ | ◯ | ◯ |

**30. In your opinion, about how many patients could be tested with this test in an 8-hour day?** *Mark only one oval per row.*

◯ < 10
◯ 10 – 50
◯ 50 – 100
◯ > 100

**31. Please comment here if you see any potential issues or room for improvement.**

*<u>READ-OUT of TEST</u>*

**32. How did you find the results read-out in the following areas:**
*Mark only one oval per row.*

| | Very easy | Easy | Neither | Difficult | Very difficult |
|---|---|---|---|---|---|
| a) Visibility of the control (C) and in contrast with the background (if applicable)? | ◯ | ◯ | ◯ | ◯ | ◯ |
| b) Visibility of the test (T) band in contrast with the background (if applicable)? | ◯ | ◯ | ◯ | ◯ | ◯ |
| c) Read-out from Reader (if applicable) | ◯ | ◯ | ◯ | ◯ | ◯ |
| d) Interpretation of the test result | ◯ | ◯ | ◯ | ◯ | ◯ |

**33. Do you foresee any issues with reading these results considering the lighting conditions in the settings you currently work or have experience with?** *Mark only one oval.*

◯ Yes (please explain below)

◯ No

**If yes, please explain here**

**34. For visual readout: Was there any color on the background of the test result (T) or control band (C) that made the interpretation of the bands difficult?** *Mark only one oval.*

◯ Not applicable
◯ Yes (please explain below)
◯ No

**If yes, which background color was present?**

**35. How satisfied are you with the reader if applicable (in terms of ease of use and fit for purpose)?**

◯ Not applicable

*If applicable, mark only one oval.*

| | 1 | 2 | 3 | 4 | 5 | |
|---|---|---|---|---|---|---|
| Very satisfied | ◯ | ◯ | ◯ | ◯ | ◯ | Very dissatisfied |

**36. Overall, how satisfied are you with the reader (if applicable)?**

◯ Not applicable

*If applicable, mark only one oval.*

| | 1 | 2 | 3 | 4 | 5 | |
|---|---|---|---|---|---|---|
| Very satisfied | ◯ | ◯ | ◯ | ◯ | ◯ | Very dissatisfied |

**37. Which component(s) of the reader (if applicable) should be improved in your opinion (if any)?**
*Please specify why and how*

*OVERALL ASSESMMENT*

**38. Overall, how did you find the use of this rapid COVID-19 diagnostic tool:** *Mark only one oval.*

| | 1 | 2 | 3 | 4 | 5 | |
|---|---|---|---|---|---|---|
| Very easy | ◯ | ◯ | ◯ | ◯ | ◯ | Very difficult |

**39. Please comment here on the use:**

---

**40. Which option(s) do you consider feasible in your setting?** *Tick all that apply.*

- ◯ Sequential testing (run tests one by one)
- ◯ Batch testing (run multiple tests at the time)

***41.*** **Which aspect(s) of this test could cause difficulties in its day-to-day use?** *Tick all that apply.*

- ◯ Hands-on time
- ◯ Tatal assay time to result
- ◯ Batch processing
- ◯ Throughput
- ◯ Test results interpretation
- ◯ Overall number of steps
- ◯ Time sensitive steps
- ◯ Cartridge design
- ◯ Quality of material
- ◯ Training requirements
- ◯ Storage conditions and stability
- ◯ Waste management requirements
- ◯ I don’t know
- ◯ None, I see no barriers for implementation

**42. Please give a short explanation for each of the aspects you selected above e.g. what could be the challenges in the day-to-day use:**

---

*SETTINGS UF USE*

**43. Do you see this test being used in its current form in your setting in your country?** *Mark only one oval*

- Yes (please explain below)
- No (please explain below)
- I don't know

Please elaborate:

---

**44. If yes, at which health care level(s) do you see this test being implemented in your country**
*Tick all that apply.*

- Family doctor / General physician
- Peripheral hospital / lab
- Reference hospital / lab
- At a testing site operated by traines staff without specific laboratory expertise

**45. If you don't see this test being used in its current form, which aspects should be changed to make it suitable for use in your setting in your country:**

---

**46. Do you see this test being used in its current form in your setting in LOW and MIDDLE INCOME COUNTRIES?**

- Yes (please explain below)
- No (please explain below)
- I don't know

Please elaborate:

---

**47. At which health care level(s) do you see this test being implemented in low and middle income countries?**

- Family doctor / General physician
- Primary health care
- Health centre / microscopy lab
- District hospital / lab
- Reference hospital / lab
- Other:

I cannot answer this questions as I have no work experience in those countris

**48.** **If you don't see this test being used in its current form, which aspects should be changed to make it suitable for use in your setting in low and middle income countries?**

**49.** **Anything else you would like to add?**

**THANK YOU VERY MUCH!**

### (E) Figure 1: Matrix for Ease of Use Assessment

| # | Question | green | green | green | yellow | yellow | amber | amber |
|---|---|---|---|---|---|---|---|---|
| 1 | User Identifier (first name, surname) | - | - | - | - | - | - | - |
| 2 | Date of filling the questionnaire | - | - | - | - | - | - | - |
| 3 | In which country do you currently work? | - | - | - | - | - | - | - |
| 4 | At which facility / study site do you currently work? | - | - | - | - | - | - | - |
| 5 | Which test are you assessing? | - | - | - | - | - | - | - |
| 6 | About how many times did you perform this test approximately? | - | - | - | - | - | - | - |
| 7 | About how many times did you observe the use of this test (not performed yourself)? | - | - | - | - | - | - | - |
| 8 | What is your profession? | - | - | - | - | - | - | - |
| 9 | How many years of laboratory experience do you have? | - | - | - | - | - | - | - |
| 10 | How many years of working experience in limited resource settings do you have? | - | - | - | - | - | - | - |
| 11 | How much experience do you have with interpreting the results of lateral flow tests or rapid diagnostics (e.g. for HIV, malaria, pregnancy)? | - | - | - | - | - | - | - |
| 12 | How satisfied were you with the following components of the test training? [Instructions for use] | very satisfied | satisfied | - | neither | - | dissatisfied | very dissatisfied |
| 12 | How satisfied were you with the following components of the test training? [Standard operating procedures] | - | - | - | - | - | - | - |
| 12 | How satisfied were you with the following components of the test training? [Face - to - face demonstration] | - | - | - | - | - | - | - |
| 13 | What additional materials (if any) do you think should be provided as part of the training? | None | - | - | 1-2 | - | >2 | - |
| 14 | How long should the training of this test be? | Self - explanatory, no | 1 - 2 hours of training | 2 - 4 hours of training | Half a day | - | Full Day | - |
| 15 | Do you consider proficiency testing necessary? | No | - | - | - | - | Yes | - |
| 16 | Please comment here on the need for proficiency testing: | - | - | - | - | - | - | - |
| 17 | After how many of these tests do you feel you could perform the test on your own? | 1 - 2 tests | - | - | 3 - 5 tests | - | 6 - 10 tests | 10 tests |
| 18 | How satisfied are you with the quality of each of the components in the kit (in terms of ease of use and fit for purpose)? [External paper box of the kit] | very satisfied | satisfied | - | neither | - | dissatisfied | very dissatisfied |
| 18 | How satisfied are you with the quality of each of the components in the kit (in terms of ease of use and fit for purpose)? [Assay diluent tube] | very satisfied | satisfied | - | neither | - | dissatisfied | very dissatisfied |
| 18 | How satisfied are you with the quality of each of the components in the kit (in terms of ease of use and fit for purpose)? [Filter cap (if necessary)] | very satisfied | satisfied | - | neither | - | dissatisfied | very dissatisfied |
| 18 | How satisfied are you with the quality of each of the components in the kit (in terms of ease of use and fit for purpose)? [Swab for specimen collection] | very satisfied | satisfied | - | neither | - | dissatisfied | very dissatisfied |
| 18 | How satisfied are you with the quality of each of the components in the kit (in terms of ease of use and fit for purpose)? [Test cartridge - device] | very satisfied | satisfied | - | neither | - | dissatisfied | very dissatisfied |
| 18 | How satisfied are you with the quality of each of the components in the kit (in terms of ease of use and fit for purpose)? [Test cartridge - packing pouch] | very satisfied | satisfied | - | neither | - | dissatisfied | very dissatisfied |
| 18 | How satisfied are you with the quality of each of the components in the kit (in terms of ease of use and fit for purpose)? [Reader (if necessary)] | very satisfied | satisfied | - | neither | - | dissatisfied | very dissatisfied |
| 19 | Which kit component(s) should be improved in your opinion (if any)? | - | - | - | - | - | - | - |
| 20 | Overall, how satisfied are you with the kit components? | 1 | 2 | - | 3 | - | 4 | 5 |
| 21 | How satisfied are you with the design of the following features? [Size of the cartridge] | very satisfied | satisfied | - | neither | - | dissatisfied | very dissatisfied |
| 21 | How satisfied are you with the design of the following features? [Space for labeling on the front (e.g. patient ID)] | very satisfied | satisfied | - | neither | - | dissatisfied | very dissatisfied |
| 21 | How satisfied are you with the design of the following features? [Size of the well to add sample mix] | very satisfied | satisfied | - | neither | - | dissatisfied | very dissatisfied |
| 21 | How satisfied are you with the design of the following features? [Size of reading window] | very satisfied | satisfied | - | neither | - | dissatisfied | very dissatisfied |
| 21 | How satisfied are you with the design of the following features? [Logical sequence of steps] | - | - | - | - | - | - | - |
| 22 | Overall, how satisfied are you with the design of the device? | 1 | 2 | - | 3 | - | 4 | 5 |
| 23 | Which part(s) of the device could be improved in your opinion (if any)? | - | - | - | - | - | - | - |
| 24 | Please assess the test's storage conditions. [Stability of test] | 12 months | - | - | 12 to 6 months | 5 to 3 months | 3 months | - |
| 24 | Please assess the test's storage conditions. [Stability of control material (if applicable)] | 12 months | - | - | 12 to 6 months | 5 to 3 months | 3 months | - |
| 24 | Please assess the test's storage conditions. [Storage temperature] | 2 - 40° | - | - | 15- 35° | 15 - 30° | 20 - 25° | - |
| 25 | Overall, how satisfied are you with the test's storage conditions? | 1 | 2 | - | 3 | - | 4 | 5 |
| 26 | Please determine the difficulty of the following steps: [a) Check expiry date] | Very easy | Easy | - | Neither | - | Difficult | Very difficult |
| 26 | Please determine the difficulty of the following steps: [b) Remove the test cartridge from the pouch] | Very easy | Easy | - | Neither | - | Difficult | Very difficult |
| 26 | Please determine the difficulty of the following steps: [c) Label the test cartridge with the patient identifier] | Very easy | Easy | - | Neither | - | Difficult | Very difficult |
| 26 | Please determine the difficulty of the following steps: [d) Label the assay diluent tube with the patient identifier] | Very easy | Easy | - | Neither | - | Difficult | Very difficult |
| 26 | Please determine the difficulty of the following steps: [e) Open the assay diluent tube by removing the seal (if appl.)] | Very easy | Easy | - | Neither | - | Difficult | Very difficult |
| 26 | Please determine the difficulty of the following steps: [f) Transfer of buffer into diluent tube (if appl.)] | Very easy | Easy | - | Neither | - | Difficult | Very difficult |
| 26 | Please determine the difficulty of the following steps: [g) Insert the swab into the tube] | Very easy | Easy | - | Neither | - | Difficult | Very difficult |
| 26 | Please determine the difficulty of the following steps: [h) Ease of swab extraction procedure] | Very easy | Easy | - | Neither | - | Difficult | Very difficult |
| 26 | Please determine the difficulty of the following steps: [i) Ability to perform extraction procedure consistently] | Very easy | Easy | - | Neither | - | Difficult | Very difficult |
| 26 | Please determine the difficulty of the following steps: [j) Ability to maintain cleanliness of ancillary devices (e.g. pipette) in order to avoid cross-contamination] | Very easy | Easy | - | Neither | - | Difficult | Very difficult |
| 26 | Please determine the difficulty of the following steps: [k) Ease of transferring sample onto device] | Very easy | Easy | - | Neither | - | Difficult | Very difficult |
| 26 | Please determine the difficulty of the following steps: [l) Ease of transferring exact quantity into the sample well] | Very easy | Easy | - | Neither | - | Difficult | Very difficult |
| 26 | Please determine the difficulty of the following steps: [m) Trouble shooting] | Very easy | Easy | - | Neither | - | Difficult | Very difficult |
| 27 | How satisfied are you with the logical sequence of steps? | 1 | 2 | - | 3 | - | 4 | |
| 28 | Overall, how difficult did you find the steps? | 1 | 2 | - | 3 | - | 4 | 4 |
| 29 | Please assess the time relevant components of the test. [Pre-analytic time] | 2 min | - | - | 3 to 5 min | 6 to 10 min | 10 min | - |
| 29 | Please assess the time relevant components of the test. [Analytic time] | 2 min | - | - | 3 to 5 min | 6 to 10 min | 10 min | - |
| 30 | In your opinion, about how many patients could be tested with this test in an 8-hour day? | 100 | - | - | 50 - 100 | - | 10 - 50 | 10 |
| 31 | Please comment here if you see any potential issues or room for improvement. | - | - | - | - | - | - | - |
| 32 | How did you find the results read-out in the following areas: [a) Visibility of the control (C) band in contrast with the background (if appl.)] | Very easy | Easy | - | Neither | - | Difficult | Very difficult |
| 32 | How did you find the results read-out in the following areas: [b) Visibility of the test (T) band in contrast with the background (if appl.)] | Very easy | Easy | - | Neither | - | Difficult | Very difficult |
| 32 | How did you find the results read-out in the following areas: [c) Read-out from Reader (if appl.)] | Very easy | Easy | - | Neither | - | Difficult | Very difficult |
| 32 | How did you find the results read-out in the following areas: [d) Interpretation of the test result] | Very easy | Easy | - | Neither | - | Difficult | Very difficult |
| 33 | Do you foresee any issues with reading these results considering the lighting conditions in the settings you currently work or have experience with? | - | - | - | - | - | - | - |
| 33 | Please elaborate | - | - | - | - | - | - | - |
| 34 | For visual readout: Was there any color on the background of the test result (T) or control band (C) that made the interpretation of the bands difficult? | - | - | - | - | - | - | - |
| 34 | If yes, which background color was present | - | - | - | - | - | - | - |
| 35 | How satisfied are you with the reader (if applicable) in terms of ease of use and fit for purpose? | 1 | 2 | - | 3 | - | 4 | 5 |
| 36 | Overall, how satisfied are you with the reader (if applicable)? | 1 | 2 | - | 3 | - | 4 | 5 |
| 37 | Which component(s) of the reader (if applicable) should be improved in your opinion (if any)? | - | - | - | - | - | - | - |
| 38 | Overall, how did you find the use of this rapid COVID-19 diagnostic tool? | 1 | 2 | - | 3 | - | 4 | 5 |
| 39 | Please comment here on the use: | - | - | - | - | - | - | - |
| 40 | Which option(s) do you consider feasible in your setting? | Batch testing (run mul | - | - | - | - | Sequential testing (rur | - |
| 41 | Which aspect(s) of this test could cause difficulties in its day-to-day use? | 0 | - | - | 1 | 2 | 3 | 4 |
| 42 | Please give a short explanation for each of the aspects you selected above e.g. what could be the challenges in the day-to-day use: | - | - | - | - | - | - | - |
| 43 | Do you see this test being used in its current form in your setting in your country? | Yes (please explain below) | | - | - | - | No (please explain bel | - |
| 43 | Please elaborate | - | - | - | - | - | - | - |
| 44 | If yes, at which health care level(s) do you see this test being implemented in your country? | At a testing site opera | Family doctor / Gener | - | Peripheral hospital / la | - | Reference hospital / la | - |
| 45 | If you don't see this test being used in its current form, which aspects should be changed to make it suitable for use in your setting in your country: | none | - | - | 1-2 | - | >2 | - |
| 46 | Do you see this test being used in its current form in your setting in LOW and MIDDLE INCOME COUNTRIES? | Yes (please explain be | - | - | - | - | No (please explain bel | - |
| 46 | Please elaborate | - | - | - | - | - | - | - |
| 47 | At which health care level(s) do you see this test being implemented in low and middle income countries? | Primary health care | Famildy doctor / Gene | Health centre / micros | District hospital / labo | - | Reference hospital / la | - |
| 48 | If you don't see this test being used in its current form, which aspects should be changed to make it suitable for use in your setting in low and middle income countries? | none | - | - | 1 | 2 | 3 | 4 |
| 49 | Anything else you would like to add? | - | - | - | - | - | - | - |

#### (F) Table 2: Study population characteristics by study site: Bioeasy

| | Overall | Heidelberg | Berlin |
|---|---|---|---|
| **Age** - Information available for N=727 | | | |
| Mean<br>Standard Deviation | 42·7<br>14·9 | 44·2<br>15·4 | 39·5<br>13·2 |
| **Gender –** Information available for N= 697 | | | |
| Women | 368<br>(52·8%) | 271<br>(38·9%) | 97<br>(13·9%) |
| Men | 329<br>(47·2%) | 203<br>(29·1%) | 126<br>(18·1%) |
| **Data combined: Overweight > BMI 25** – Information available on N=688 | | | |
| Yes | 247<br>(36·4%) | 162<br>(23·9%) | 85<br>(12·5%) |
| No | 436<br>(63·6%) | 302<br>(44·5%) | 130<br>(19·1%) |
| **Comorbidities** – Information available on N=727 | | | |
| All with comorbidities | 304<br>(41·8%) | 231<br>(31·8%) | 73<br>(10·0%) |
| Lung diseases | | | |
| Asthma bronchiale | 57<br>(7·8%) | 41<br>(5·6%) | 16<br>(2·2%) |
| Chronic obstructive pulmonary disease (COPD) | 8<br>(1·1%) | 7<br>(1·0%) | 1<br>(0·1%) |
| Breathing disorders during sleep and obstructive Sleep Apnea | 19<br>(2·6%) | 19<br>(2·6%) | 0 |
| Interstitial Lung Disease | 2<br>(0·3%) | 2<br>(0·3%) | 0 |
| Lung Cancer | 1<br>(0·1%) | 1<br>(0·1%) | 0 |
| Other | 19<br>(2·6%) | 14<br>(1·9%) | 5<br>(0·7%) |
| Other diseases | | | |
| Cardiovascular diseases | 123<br>(18·8%) | 99<br>(15·1%) | 24<br>(3·7%) |
| Chronic kidney diseases | 9<br>(1·4%) | 7<br>(1·1%) | 2<br>(0·3%) |
| Autoimmune | 46<br>(7·2%) | 34<br>(5·3%) | 12<br>(1·9%) |
| HIV | 1<br>(0·2%) | 0 | 1<br>(0·2%) |
| Others | 143<br>(19·9%) | 110<br>(15·3%) | 33<br>(4·6%) |
| **Previous tested negative** – Information available for N=623 | | | |
| Yes<br><br>No | 73<br>(11·7%)<br>550<br>(88.3%) | 51<br>(8·2%)<br>383<br>(61·5%) | 22<br>(3·5%)<br>167<br>(26·8%) |

| | | | |
|---|---|---|---|
| **Symptoms on testing day** – Information available for N=695 | | | |
| Yes<br><br>No | 564<br>(81·2%)<br>131<br>(18·8%) | 367<br>(52·8%)<br>107<br>(15·4%) | 197<br>(28·3%)<br>24<br>(3·5%) |
| List of symptoms reported | | | |
| Fever | 189<br>(32·9%) | 147<br>(25·6%) | 42<br>(7·3%) |
| Fever measured<br>1. <38.4<br>2. >38.5 and <=39.4<br>3. >39.5 and >40.5 | <br>114 (69·5%)<br>41 (25·0%)<br>9 (5·5%) | <br>87 (53·0%)<br>35(21·3%)<br>7 (4·3%) | <br>27 (16·5%)<br>6 (3·7%)<br>2 (1·2%) |
| Cough | 318<br>(54·0%) | 219<br>(37·2%) | 99<br>(16·8%) |
| Productive cough | 101<br>(17·8%) | 76<br>(13·4%) | 25<br>(4·4%) |
| Sore throat | 312<br>(53·1%) | 191<br>(32·5%) | 121<br>(20·6%) |
| Shortness of breath | 92<br>(16·0%) | 83<br>(14·5%) | 9<br>(1·6%) |
| Muscle pain | 209<br>(36·3%) | 156<br>(27·1%) | 53<br>(9·2%) |
| Fatigue | 385<br>(65·4%) | 271<br>(46·0%) | 114<br>(19·4%) |
| Headache | 315<br>(53·6%) | 216<br>(36·7%) | 99<br>(16·8%) |
| Runny nose | 188<br>(32·8%) | 131<br>(22·8%) | 57<br>(9·9%) |
| Chest pain | 98<br>(17·3%) | 88<br>(15·5%) | 10<br>(1·8%) |
| Diarrhea | 93<br>(16·3%) | 69<br>(12·1%) | 24<br>(4·2%) |
| Nausea | 59<br>(10·5%) | 44<br>(7·8%) | 15<br>(2·7%) |
| Loss of taste and smell | 63<br>(11·1%) | 46<br>(8·1%) | 17<br>(3·0%) |
| Others | 105<br>(21·0%) | 73<br>(14·6%) | 32<br>(6·4%) |

#### **(G)** Table 3: Study population characteristics by study site: Coris

| | **Overall** | **Heidelberg** | **Berlin** | **Liverpool** |
|---|---|---|---|---|
| **Age** - Information available for N= 416 | | | | |
| Mean<br>Standard Deviation | 45·1<br>15·4 | 46·4<br>14·5 | * | 64·3<br>11·7 |
| **Gender** – Information available for N= 411 | | | | |
| Women | 248<br>(60·3%) | 175<br>(42·6%) | 56<br>(13·6%) | 17<br>(4·1%) |
| Men | 162<br>(39·4%) | 99<br>(24·1%) | 47<br>(11·4%) | 16<br>(3·9%) |

| | | | | |
|---|---|---|---|---|
| Diverse | 1 (0·2%) | 1 (0·2%) | 0 | 0 |
| **Data combined: Overweight > BMI 25** – Information available on N= 380 | | | | |
| Yes | 181 (47·6%) | 156 (41·1%) | 20 (5·3%) | 5 (1·3%) |
| No | 199 (52·4%) | 119 (31·3%) | 54 (14·2%) | 26 (6·8%) |
| **Comorbidities** – Information available on N= 417 | | | | |
| All comorbidities | 198 (47·5%) | 142 (34·1%) | 24 (5·8%) | 32 (7·7%) |
| | Lung diseases | | | |
| Asthma bronchiale | 41 (9·8%) | 25 (6·0%) | 8 (1·9%) | 8 (1·9%) |
| Chronic obstructive pulmonary disease (COPD) | 23 (5·5%) | 4 (1·0%) | 2 (0.5%) | 17 (4·1%) |
| Breathing disorders during sleep and Obstructive Sleep Apnea | 10 (2·4%) | 7 (1·7%) | 1 (0·2%) | 2 (0·5%) |
| Interstitial Lung Disease | 0 | 0 | 0 | 0 |
| Lung Cancer | 1 (0·2%) | 0 | 0 | 1 (0·2%) |
| Other | 11 (2·6%) | 11 (2·6%) | 0 | 0 |
| | Other diseases | | | |
| Cardiovascular diseases | 71 (17·7%) | 48 (11·9%) | 4 (1·0%) | 19 (4·7%) |
| Chronic kidney diseases | 11 (2·7%) | 7 (1·7%) | 0 | 4 (1·0%) |
| Autoimmune | 29 (7·3%) | 24 (6·0%) | 2 (0·5%) | 3 (0·8%) |
| HIV | 0 | 0 | 0 | 0 |
| Others | 109 (26·1%) | 74 (17·7%) | 10 (2·4%) | 25 (6·0%) |
| **Previous tested negative** – Information available for N= 302 | | | | |
| Yes<br>No | 38 (12·6%)<br>264 (87·4%) | 28 (9·3%)<br>214 (70·9%) | 10 (3·3%)<br>50 (16·6%) | NA |
| **Symptoms on testing day** – Information available for N= 411 | | | | |
| Yes<br>No | 283 (68·9%)<br>128 (31·1%) | 149 (36·3%)<br>127 (30·9%) | 101 (24·6%)<br>1 (0·2%) | 33 (8·0%)<br>0 |
| | List of symptoms reported | | | |
| Fever | 84 (30·9%) | 52 (19·1%) | 20 (7·4%) | 12 (4·4%) |
| Fever measured<br>1. <38.4 | 39 (61·9%) | 30 (47·6%) | 9 | NA |

| | | | | |
|---|---|---|---|---|
| 2. >38.5 and <=39.4<br>3. >39.5 and >40.5 | 20 (31·7%)<br>4 (6·3%) | 13 (20·6%)<br>4 (6·3%) | 7<br>0 | |
| Cough | 155 (55·8%) | 77 (27·7%) | 53 (19·1%) | 25 (9·0%) |
| Productive cough | 44 (17·6%) | 24 (9·6%) | 8 (3·2%) | 12 (4·8%) |
| Sore throat | 162 (58·5%) | 88 (31·8%) | 74 (26·7%) | 0 |
| Shortness of breath | 60 (22·2%) | 26 (9.6%) | 4 (1·5%) | 30 (11·1%) |
| Muscle pain | 108 (39·3%) | 68 (24·7%) | 38 (13·8%) | 2 (0·7%) |
| Fatigue | 183 (66·1%) | 103 (70.3%) | 71 (25·6%) | 9 (3·2%) |
| Headache | 154 (55·4%) | 96 (34·5%) | 57 (20·5%) | 1 (0·4%) |
| Runny nose | 102 (37·4%) | 50 (18·3%) | 52 (19·0%) | 0 |
| Chest pain | 44 (16·3%) | 32 (11·9%) | 2 (0·7%) | 10 (3·7%) |
| Diarrhea | 43 (15·8%) | 30 (11·0%) | 11 (4·0%) | 2 (0·7%) |
| Nausea | 36 (14·7%) | 24 (9·8%) | 4 (1·6%) | 8 (3·3% |
| Loss of taste and smell | 30 (11·1%) | 18 (6·6%) | 10 (3·7%) | 2 (0·7%) |
| Others | 48 (18·4%) | 28 (10·7%) | 5 (1·9%) | 15 (5·7%) |

#### (H) Table 4: Study population characteristics by study site: SD Biosensor

| | **Overall** | **Heidelberg** | **Berlin** | **Liverpool** |
|---|---|---|---|---|
| **Age** - Information available for N=1262 | | | | |
| Mean<br>Standard Deviation | 37·6<br>12·7 | 40·9<br>14·0 | 35·8<br>11·3 | 66·1<br>11·3 |
| **Gender** – Information available for N= 1254 | | | | |
| Women | 630 (50·2%) | 188 (15·0%) | 435 (34·7%) | 7 (0·6%) |
| Men | 624 (49·8%) | 138 (11·0%) | 475 (37·9%) | 11 (0·9%) |
| **Data combined: Overweight > BMI 25** – Information available on N=1197 | | | | |
| Yes | 451 (36·9%) | 145 (11·9%) | 305 (25·0%) | 1 (0·1%) |
| No | 771 (63·1%) | 181 (14·8%) | 575 (47·1%) | 15 (1·2%) |
| **Comorbidities** – Information available on N=1263 | | | | |

| | | | | |
|---|---|---|---|---|
| All with comorbidities | 361 (28·6%) | 125 (9·9%) | 218 (17·3%) | 18 (1·4%) |
| Lung diseases | | | | |
| Asthma bronchiale | 83 (6·6%) | 29 (2·3%) | 51 (4·0%) | 3 (0·2%) |
| Chronic obstructive pulmonary disease (COPD) | 18 (1·4%) | 3 (0·2%) | 5 (0·4%) | 10 (0·8%) |
| Breathing disorders during sleep and obstructive Sleep Apnea | 13 (1·0%) | 10 (0·8%) | 2 (0·2%) | 1 (0·1%) |
| Interstitial Lung Disease | 1 (0.1%) | 0 | 1 (0.1%) | 0 |
| Lung Cancer | 2 (0·2%) | 0 | 1 (0·1%) | 1 (0·1%) |
| Other | 30 (2·4%) | 5 (0·4%) | 25 (2·0%) | 0 |
| Other diseases | | | | |
| Cardiovascular diseases | 108 (9·3%) | 37 (3·2%) | 61 (5·2%) | 10 (0·9%) |
| Chronic kidney diseases | 11 (0·9%) | 6 (0·5%) | 4 (0·3%) | 1 (0·1%) |
| Autoimmune | 42 (3·6%) | 21 (1·8%) | 19 (1·6%) | 2 (0·2%) |
| HIV | 5 (0·4%) | 0 | 5 (0·4%) | 0 |
| Others | 136 (10·8%) | 52 (4·1%%) | 70 (5·5%) | 14 (1·1%) |
| **Previous tested negative** – Information available for N=1003 | | | | |
| Yes<br>No | 125 (12·5%)<br>878 (87·5%) | 39 (3·9%)<br>242 (24·1%) | 86 (8·6%)<br>636 (63·4%) | NA |
| **Symptoms on testing day** – Information available for N=1249 | | | | |
| Yes<br>No | 1054 (84·4%)<br>195 (15·6%) | 204 (16·3%)<br>122 (9·8%) | 832 (66·6%)<br>73 (5·8%) | 18 (1·4%)<br>0 |
| List of symptoms reported | | | | |
| Fever | 190 (20·5%) | 42 (4·5%) | 141 (15·2%) | 7 (0·8%) |
| Fever measured<br>1. <38.4<br>2. >38.5 and <=39.4<br>3. >39.5 and >40.5 | <br>96 (68·5%)<br>37 (26·4%)<br>7 (5·0%) | <br>26 (18·6%)<br>7 (5·0%)<br>4 (2·7%) | <br>70 (50·0%)<br>30 (21·4%)<br>3 (2·1%) | NA |
| Cough | 528 (54·1%) | 115 (11·8%) | 398 (40·8%) | 15 (1·5%) |
| Productive cough | 141 (18·5%) | 48 (6·3%) | 85 (11·2%) | 8 (1·1%) |
| Sore throat | 655 (66·4%) | 136 (13·8%) | 519 (52·6%) | 0 |
| Shortness of breath | 52 (5·7%) | 14 (1·5%) | 22 (2·4%) | 16 (1·8%) |
| Muscle pain | 319 (34·2%) | 72 (7·7%) | 246 (26·4%) | 1 (0·1%) |

| | | | | |
|---|---|---|---|---|
| Fatigue | 663 (68·1%) | 152 (15·6%) | 505 (51·8%) | 6 (0·6%) |
| Headache | 547 (56·7%) | 126 (13·1%) | 420 (43·7%) | 1 (0·1%) |
| Runny nose | 451 (48·1%) | 115 (12·3%) | 336 (35·8%) | 0 |
| Chest pain | 43 (4·8%) | 20 (2·2%) | 17 (1·9%) | 6 (0·7%) |
| Diarrhea | 127 (13·8%) | 28 (3·0%) | 97 (10·6%) | 2 (0·1%) |
| Nausea | 66 (8·7%) | 14 (1·8%) | 46 (6·0%) | 6 (0·8%) |
| Loss of taste and smell | 106 (11·7%) | 24 (2·6%) | 81 (8·9%) | 1 (0·1%) |
| Others | 119 (17·4%) | 21 (3·1%) | 90 (13·1%) | 8 (1·2%) |

#### (I) Table 5: Detailed list of symptoms for all PCR positives

| | Ag test result | Fever | Cough | Expectorate cough | Sore throat | Shortness of breath | Muscle pain | Fatigue | Headache | Runny nose | Chest pain | Diarrhea | Nausea | Loss of taste or smell | Other |
|---|---|---|---|---|---|---|---|---|---|---|---|---|---|---|---|
| 1 | negative | 0 | 1 | 0 | 1 | 0 | 1 | 1 | 1 | 1 | 1 | 1 | 1 | | 1 |
| 2 | negative | 0 | 1 | 0 | 0 | 0 | 0 | 1 | 1 | 0 | 0 | 0 | 0 | 1 | 0 |
| 3 | negative | NA | NA | NA | NA | NA | NA | NA | NA | NA | NA | NA | NA | NA | NA |
| 4 | negative | 1 | 0 | 0 | 0 | 0 | 1 | 1 | 1 | 0 | 0 | 0 | 1 | 1 | 0 |
| 5 | negative | 0 | 1 | 0 | 1 | 0 | 0 | 1 | 0 | 0 | 0 | 1 | 0 | 0 | 0 |
| 6 | positive | 0 | 0 | 0 | 0 | 0 | 1 | 1 | 0 | 0 | 0 | 0 | 0 | 1 | |
| 7 | positive | 0 | 0 | 0 | 0 | 0 | 0 | 1 | 1 | 1 | 1 | 0 | 0 | 1 | 0 |
| 8 | positive | 0 | 0 | 0 | 0 | 0 | 0 | 1 | 0 | 0 | 0 | 0 | 0 | 1 | 0 |
| 9 | positive | 0 | 1 | 0 | 1 | 0 | 1 | 1 | 1 | 0 | 0 | 0 | 0 | 0 | 0 |
| 10 | positive | 0 | 0 | 0 | 0 | 0 | 1 | 1 | 0 | 0 | 0 | 0 | 0 | 1 | 0 |
| 11 | positive | NA | NA | NA | NA | NA | NA | NA | NA | NA | NA | NA | NA | NA | NA |
| 12 | negative | NA | NA | NA | NA | NA | NA | NA | NA | NA | NA | NA | NA | NA | NA |
| 13 | negative | NA | NA | NA | NA | NA | NA | NA | NA | NA | NA | NA | NA | NA | NA |
| 14 | positive | 1 | | 1 | 0 | 0 | 1 | 0 | 0 | 0 | 0 | 0 | 0 | 0 | 1 |
| 15 | positive | 0 | 1 | 0 | 0 | 0 | 0 | 0 | 0 | 0 | 0 | 0 | 0 | 0 | 0 |
| 16 | positive | NA | NA | NA | NA | NA | NA | NA | NA | NA | NA | NA | NA | NA | NA |

| | | | | | | | | | | | | | | | |
|---|---|---|---|---|---|---|---|---|---|---|---|---|---|---|---|
| 17 | positive | 0 | 0 | 0 | 0 | 0 | 0 | 0 | 1 | 0 | 0 | 0 | 0 | 1 | 0 |
| 18 | positive | 0 | 0 | 0 | 1 | 0 | 0 | 1 | 1 | 0 | 0 | 0 | 1 | 0 | 0 |
| 19 | negative | | 1 | | | | | | | | | | | | |
| 20 | negative | | | | | | | | 1 | | | | | | |
| 21 | negative | 0 | 0 | 0 | 0 | 0 | 0 | 0 | 1 | 0 | 0 | 0 | 0 | 0 | 0 |
| 22 | negative | 1 | 1 | 0 | 1 | 0 | 0 | 1 | 0 | 1 | 0 | 0 | 0 | 1 | |
| 23 | negative | 0 | 1 | 0 | 0 | 0 | 1 | 1 | 0 | 1 | 0 | 0 | 0 | 0 | 0 |
| 24 | negative | 0 | 1 | 0 | 1 | 0 | 0 | 1 | 0 | 1 | 0 | 1 | 0 | 0 | 0 |
| 25 | negative | 0 | 1 | 0 | 0 | 1 | 0 | 1 | 0 | 0 | 0 | 0 | 0 | 0 | 0 |
| 26 | negative | | 1 | 0 | | | | | 1 | | | | 0 | | |
| 27 | negative | 0 | 0 | 0 | 1 | 0 | 0 | 0 | 0 | 0 | 0 | 0 | 0 | 1 | |
| 28 | negative | 0 | 1 | | 0 | 0 | 1 | 1 | 1 | 1 | 0 | 1 | | 1 | 1 |
| 29 | negative | 1 | 0 | | 1 | 0 | 0 | 1 | 0 | 0 | 0 | 1 | | 0 | 1 |
| 30 | negative | NA | NA | NA | NA | NA | NA | NA | NA | NA | NA | NA | NA | NA | NA |
| 31 | negative | 0 | 0 | | 0 | 0 | 0 | 0 | 1 | 0 | 0 | 0 | | 0 | |
| 32 | positive | 0 | 0 | 0 | 1 | 0 | 0 | 0 | 0 | 0 | 0 | 0 | 0 | 1 | 0 |
| 33 | positive | 1 | 1 | 1 | 1 | 0 | 0 | 1 | 1 | 0 | 0 | 0 | 0 | 1 | 0 |
| 34 | positive | 1 | 1 | | | | | 1 | | | | | | 1 | |
| 35 | positive | 0 | 1 | 0 | 1 | 0 | 0 | 1 | 1 | 1 | 0 | 0 | 0 | 1 | 0 |

| | | | | | | | | | | | | | | | |
|---|---|---|---|---|---|---|---|---|---|---|---|---|---|---|---|
| 36 | positive | 0 | 0 | 0 | 1 | 0 | 0 | 0 | 1 | 0 | 0 | 0 | 0 | 0 | 0 |
| 37 | positive | 0 | 0 | 0 | 0 | 0 | 0 | 1 | 1 | 0 | 0 | 0 | 0 | 1 | 0 |
| 38 | positive | 1 | 1 | 0 | 1 | 0 | 0 | 0 | 1 | 0 | 0 | 0 | 0 | 0 | 0 |
| 39 | positive | 0 | 1 | 1 | 1 | 1 | 1 | 1 | 0 | 1 | 0 | 1 | 0 | 1 | 0 |
| 40 | positive | 1 | 1 | 0 | 0 | 0 | 0 | 1 | 1 | 0 | 0 | 1 | | 0 | |
| 41 | positive | 0 | 1 | 0 | 0 | 0 | 1 | 1 | 1 | 0 | 0 | 0 | 0 | 1 | |
| 42 | positive | 0 | 0 | | 1 | | 1 | 1 | 1 | | | | | 1 | |
| 43 | positive | 0 | 0 | 0 | 0 | 0 | 0 | 1 | 1 | 0 | 0 | 0 | 0 | 0 | 1 |
| 44 | positive | 1 | 1 | 0 | 0 | 0 | 1 | 0 | 1 | 0 | 0 | 0 | 0 | 0 | 0 |
| 45 | positive | | 1 | | | | | | 1 | | | | | 1 | |
| 46 | positive | 0 | 0 | 0 | 0 | 0 | 0 | 1 | 0 | 0 | 0 | 0 | 0 | 0 | 0 |
| 47 | positive | 1 | 1 | 0 | 1 | 0 | 0 | 1 | 0 | 1 | 0 | 0 | 0 | 0 | 0 |
| 48 | positive | 1 | 1 | 1 | 0 | 0 | 0 | 1 | 1 | 0 | | 0 | 0 | 0 | 0 |
| 49 | positive | 0 | 0 | | 0 | 0 | 0 | 0 | 0 | 0 | 0 | 1 | | 1 | |
| 50 | positive | 0 | 1 | 0 | 1 | 0 | 0 | 1 | 0 | 0 | 0 | 0 | 0 | 0 | 0 |
| 51 | positive | 0 | 1 | 0 | 1 | 0 | 0 | 0 | 0 | 0 | 0 | 0 | 0 | 0 | 0 |
| 52 | positive | | 1 | | 1 | | 1 | 1 | 1 | | | | | 1 | |
| 53 | positive | 1 | 1 | | 1 | 0 | 1 | 1 | 1 | 0 | 0 | 0 | | 0 | 1 |
| 54 | positive | 1 | 1 | | 1 | | 1 | 1 | 1 | | | | | | |

| | | | | | | | | | | | | | | | |
|---|---|---|---|---|---|---|---|---|---|---|---|---|---|---|---|
| 55 | positive | 1 | 0 | 0 | 0 | 1 | 0 | 0 | 0 | 0 | 0 | 0 | 0 | 0 | 0 |
| 56 | positive | 0 | 0 | | 0 | 0 | 0 | 0 | 0 | 1 | 0 | 0 | | 1 | |
| 57 | positive | 0 | 1 | | 1 | 0 | 0 | 0 | 0 | 0 | 0 | 0 | | 0 | |
| 58 | positive | 1 | 1 | | 0 | 0 | 1 | 1 | 1 | 1 | 0 | 1 | | 1 | |
| 59 | positive | 0 | 0 | | 0 | 0 | 0 | 0 | 0 | | 0 | 0 | | 0 | |
| 60 | positive | 0 | 0 | | 1 | 0 | 0 | 0 | 1 | 1 | 0 | 0 | | 0 | |
| 61 | positive | 0 | 1 | | 0 | 0 | 0 | 1 | 1 | 0 | 0 | 0 | 0 | 0 | 0 |
| 62 | positive | 0 | 0 | | 1 | 0 | 0 | 1 | 1 | 0 | 0 | 0 | | 0 | |
| 63 | positive | 1 | 1 | | 0 | 0 | 1 | 1 | 1 | 1 | 0 | 0 | | 0 | |
| 64 | positive | 1 | 0 | | 1 | 0 | 1 | 1 | 1 | | 0 | 1 | | 0 | 1 |
| 65 | positive | 0 | 1 | | 0 | 0 | 0 | 0 | 0 | 0 | 0 | 0 | | 0 | 0 |
| 66 | positive | 0 | 1 | | 1 | 1 | 1 | 1 | 1 | 1 | 0 | 1 | | 0 | |
| 67 | positive | 0 | 1 | 0 | 0 | 0 | 1 | 1 | 1 | 1 | 0 | 0 | 0 | 1 | 0 |
| 68 | positive | NA | NA | NA | NA | NA | NA | NA | NA | NA | NA | NA | NA | NA | NA |
| 69 | positive | 0 | 0 | | 0 | 1 | 0 | 0 | 0 | 0 | 0 | 0 | | 1 | 0 |
| 70 | positive | 0 | 1 | | 1 | 0 | 0 | 1 | 1 | 1 | 0 | 0 | | 0 | 0 |

#### (J) Table 6: Antigen-based RDT with test result, CT values and viral load for PCR positive patients in Berlin and Heidelberg

CT-value reported here represent the E-gene genome target (similar to the target described by Corman) by descending order. A conversion of CT-values for RT-PCR tests into viral-load was performed using quantified specific in vitro-transcribed RNA (Corman 2020 Eurosurveillance).

| Antigen RDT | Antigen RDT result | Ct value (E-Gene) | Viral load (log$_{10}$ RNA SARS-CoV2/swab) | PCR assay |
|---|---|---|---|---|
| **Bioeasy** Bioeasy 2019-nCoV Ag Fluorescence Rapid Test Kit | Berlin | | | |
| | positive | 19·55 | 9·39 | Roche Cobas |
| | positive | 20·29 | 9·14 | Roche Cobas |
| | positive | 22·78 | 8·33 | Roche Cobas |
| | negative | 23·09 | 8·23 | Roche Cobas |
| | positive | 23·47 | 8·10 | Roche Cobas |
| | negative | 33·05 | 4·97 | Roche Cobas |
| | Heidelberg | | | |
| | positive | 20·63 | * | TibMolBiol |
| | positive | 21·78 | * | TibMolBiol |
| | negative | 25·73 | * | TibMolBiol |
| | positive | 28·82 | * | TibMolBiol |
| | negative | 31·29 | * | TibMolBiol |
| | positive | 32·31 | * | TibMolBiol |
| | negative | 36·90 | * | TibMolBiol |
| | Heidelberg | | | |
| | positive | 17·59 | * | Seegene |
| | positive | 17·61 | * | Seegene |
| **SD Biosensor** STANDARD Q COVID-19 Ag Test | Berlin | | | |
| | positive | 18·1 | 9·69 | Roche Cobas |
| | positive | 18·38 | 9·60 | Roche Cobas |
| | positive | 19·23 | 9·35 | Roche Cobas |
| | positive | 19·4 | 9·30 | Roche Cobas |
| | positive | 20·85 | 8·87 | Roche Cobas |

| | | | |
|---|---|---|---|
| positive | 20·98 | 8·84 | Roche Cobas |
| positive | 21·31 | 8·72 | Roche Cobas |
| positive | 21·56 | 8·66 | Roche Cobas |
| positive | 21·55 | 8·65 | Roche Cobas |
| positive | 22·11 | 8·50 | Roche Cobas |
| positive | 22·88 | 8·27 | Roche Cobas |
| positive | 22·91 | 8·25 | Roche Cobas |
| positive | 23·09 | 8·23 | Roche Cobas |
| positive | 23·19 | 8·18 | Roche Cobas |
| positive | 23·31 | 8·13 | Roche Cobas |
| positive | 24·19 | 7·89 | Roche Cobas |
| positive | 24·54 | 7·78 | Roche Cobas |
| positive | 24·96 | 7·66 | Roche Cobas |
| positive | 25·28 | 7·64 | Roche Cobas |
| positive | 25·12 | 7·58 | Roche Cobas |
| positive | 25·21 | 7·56 | Roche Cobas |
| positive | 25·76 | 7·42 | Roche Cobas |
| positive | 26·37 | 7·24 | Roche Cobas |
| positive | 26·71 | 7·14 | Roche Cobas |
| positive | 26·84 | 7·10 | Roche Cobas |
| positive | 26·96 | 7·07 | Roche Cobas |
| positive | 27·42 | 6·93 | Roche Cobas |
| negative | 27·44 | 6·89 | Roche Cobas |
| negative | 27·68 | 6·85 | Roche Cobas |
| positive | 28·23 | 6·69 | Roche Cobas |
| positive | 28·44 | 6·63 | Roche Cobas |
| positive | 29·75 | 6·20 | Roche Cobas |
| positive | 30·83 | 5·88 | Roche Cobas |
| negative | 32·2 | 5·52 | Roche Cobas |
| positive | 32·35 | 5·47 | Roche Cobas |
| negative | 34·23 | 4·92 | Roche Cobas |

| | | | | |
|---|---|---|---|---|
| | negative | 34·39 | 4·81 | Roche Cobas |
| | negative | 36·65 | 4·20 | Roche Cobas |
| | negative | 37·36 | 3·99 | Roche Cobas |
| | negative | 37·6 | 3·92 | Roche Cobas |
| | Heidelberg | | | |
| | positive | 15·75 | * | TibMolBiol |
| | negative | 30·94 | * | TibMolBiol |
| | negative | 35·89 | * | TibMolBiol |
| | Heidelberg | | | |
| | * | · | * | Seegene |
| | positive | 20·44 | * | Seegene |
| | negative | 32·70 | * | Seegene |
| | * | · | * | Seegene |
| **Coris** COVID-19 Ag Respi-Strip | Berlin | | | |
| | negative | 22·22 | 8·45 | Roche Cobas |
| | positive | 23·65 | 8·02 | Roche Cobas |
| | positive | 24·31 | 7·83 | Roche Cobas |
| | negative | 29·54 | 6·26 | Roche Cobas |
| | Heidelberg | | | |
| | positive | 19·69 | * | TibMolBiol |
| | negative | 31·66 | * | TibMolBiol |

* Results will be provided with the revision
